# Predicting long-term adverse outcomes after neonatal intensive care

**DOI:** 10.64898/2026.03.26.26348580

**Authors:** Mine Öğretir, Volmar Kaipainen, Markus Leskinen, Harri Lähdesmäki, Miika Koskinen

## Abstract

Neonates requiring intensive care are at increased risk for long-term neuropsychiatric disorders. However, clinical adoption of risk prediction models remains limited when their performance lacks adequate interpretability for informed clinical decision-making. Here, we investigated whether longitudinal neonatal electronic health record (EHR) data from the first 90 days of life can support clinically meaningful interpretation of long-term risk signals for major neuropsychiatric diagnoses by age seven. In a retrospective register-based cohort of 17,655 at-risk children from an academic medical center, of whom 8.0% (1,420) received a major neuropsychiatric diagnosis during follow-up, we applied a time-aware transformer model (Self-supervised Transformer for Time-Series; STraTS) and thoroughly evaluated its predictions using three complementary interpretability approaches: perturbation-based variable importance, value-dependent effect analysis, and leave-one-out (LOO) feature attribution. STraTS achieved the highest area under the precision–recall curve (AUPRC 0.171 *±* 0.022), compared with Random Forest (0.166 *±* 0.008), logistic regression (0.151 *±* 0.007), and XGBoost (0.128 *±* 0.010). Across interpretability methods, five predictors were consistently identified: birth weight, gender, Apgar score at 1 minute, umbilical serum thyroid stimulating hormone (uS-TSH), and treatment time in hospital. Indicators of early clinical severity, including chromosomal abnormalities and neonatal cerebral-status disturbances, showed the largest risk-increasing effects. Furthermore, the model’s learned vector representations of subject-specific EHR sequences formed clinically coherent latent embeddings that reflect population heterogeneity along established perinatal risk dimensions. These findings demonstrate that combining multiple complementary interpretability methods yields stable, clinically plausible risk signals while revealing limitations that would remain undetected by any single approach, highlighting the importance of careful interpretability analysis of deep learning-based risk predictions.

**Author summary:** Infants who require intensive care after birth are more likely to develop neuropsychiatric conditions such as cerebral palsy, epilepsy, autism, or intellectual disability later in childhood. Identifying high-risk infants early could improve follow-up care, but prediction models are difficult to trust without understanding how they reach their conclusions. We used hospital records from the first 90 days of life for nearly 17,700 children to train a machine learning model that processes clinical events over time, and we applied three different methods to explain what the model learned. The model grouped children in ways that reflected known risk factors such as prematurity and severity of illness, suggesting it captures meaningful patterns beyond any single variable. Importantly, no single explanation method told the complete story: one missed rare but serious conditions because it averaged across all patients, while another produced a misleading result for gestational age because the same information was already captured by birth weight. Only by comparing methods could we detect these issues. Our key contribution is not prediction accuracy—which remains limited by the complexity of these conditions—but demonstrating that multiple complementary explanation methods are needed to produce trustworthy insights when applying machine learning to clinical data.

## Introduction

Neonatal intensive care is associated with elevated risk for long-term neuropsychiatric morbidity extending into childhood and adolescence [1]. This increased risk reflects multiple prenatal and early-life exposures, including prematurity, perinatal injury, systemic illness, maternal medication exposure during pregnancy, and complications related to intensive care. Large population-based cohort studies, meta-analyses, and longitudinal work have consistently linked preterm birth and low birth weight to cerebral palsy, epilepsy, intellectual disability, autism spectrum disorder, and broader neurodevelopmental impairment, with effects persisting across the lifespan [2–7].

Neonatal complications, including perinatal brain injury and seizures, contribute additional independent risk for long-term neurodevelopmental impairment [8, 9]. Autism spectrum disorder has similarly been associated with preterm birth and neonatal intensive care exposure, with broader reviews confirming the relevance of early-life health trajectories for later neuropsychiatric outcomes [10, 11]. While these studies have established robust associations, they also reveal substantial heterogeneity in individual risks, underscoring the need for predictive models that can leverage longitudinal neonatal health data to capture complex, time-dependent risk patterns. This motivates the use of time-aware architectures that can model the full longitudinal neonatal electronic health record (EHR), combined with systematic interpretability analyses, to improve individual-level risk prediction and yield clinically plausible explanations of model prediction.

However, achieving this goal remains challenging. Existing approaches typically rely on time-fixed perinatal variables or coarse summary measures, which only partially capture the complexity of neonatal care trajectories. In neonatal intensive care settings, patient histories unfold as irregular, sparse, and high-dimensional longitudinal sequences comprising heterogeneous data types such as diagnoses, medication administrations, laboratory measurements, and healthcare utilization events, where the timing, accumulation, and interaction of events are clinically meaningful yet difficult to represent with static features [12]. Sequence models have been developed to learn predictive representations from such clinical time series [13], with more recent work emphasizing time-aware representations for irregularly sampled records [14]. Building on these directions, transformer-based approaches operate directly on sparse and irregular clinical event streams, reducing the need for heavy discretization and imputation [15–17]. Deep learning applied to neonatal EHRs has shown value for predicting near-term morbidity from linked longitudinal records [18], and transformer-based models have been explored for early identification of autism spectrum disorder from administrative data [19]. However, application of sequence-based models to long-term neuropsychiatric outcome prediction in neonatal intensive care populations remains limited [20], and questions of model interpretability and clinical relevance are critical for practical adoption.

For clinical prediction models intended to inform follow-up or resource allocation, interpretability is important for assessing plausibility, identifying potential spurious correlations, and supporting transparent communication of model prediction in high-dimensional EHR settings [21, 22]. Interpretability methods span global approaches that summarize model reliance on variables across the population and local approaches that explain individual predictions. Perturbation-based analyses, such as ablation or permutation of variables, provide an intuitive global view by quantifying performance changes when information from a variable is removed or disrupted [23]. Partial dependence–style analyses offer a complementary perspective by examining how model predictions vary with a variable’s value while averaging over remaining covariates [24]. Local attribution methods—such as leave-one-out (LOO) feature attribution, which evaluates prediction changes when individual features are removed—further provide instance-level explanations that complement global summaries.

In this study, we apply an established time-aware sequence modeling framework, Self-supervised Transformer for Time-Series (STraTS) [15], to predict long-term neuropsychiatric outcomes from longitudinal and time-fixed neonatal EHR data in the early postnatal period, together with maternal medication exposure during pregnancy. Our objective is to assess the utility of sequence-based representation learning for neonatal risk prediction in a real-world clinical dataset and to translate model prediction into clinically interpretable findings. Specifically, we (1) benchmark STraTS against commonly used non-sequential baselines and evaluate discrimination under class imbalance; (2) conduct complementary interpretability analyses: global variable importance via perturbation (ablation or permutation depending on variable type) and per-patient LOO feature attribution as two independent approaches for identifying influential variables, together with a value-dependent variable analysis that characterizes the direction and magnitude of prediction changes for perturbation-selected variables; and (3) examine the model’s learned patient representations by extracting time-dependent embeddings and relating them to clinically meaningful characteristics and model outputs, as a qualitative assessment of whether clinically plausible structure emerges in the learned embedding space.

Central to our approach is the combination of complementary interpretability methods that serve distinct roles. Two methods independently identify influential variables: perturbation-based importance (ablation or permutation) quantifies *predictive reliance* by measuring performance degradation when information carried by a feature is removed, providing a robust global ranking; and LOO feature attribution provides *instance-level* attributions that reveal heterogeneity and context dependence, while its aggregation yields an independent cohort-level ranking. A third analysis, value-dependent variable effects, complements the perturbation results by characterizing the *direction* and *magnitude* of prediction changes for perturbation-selected variables across realistic value ranges, indicating whether higher values (or presence) tend to increase or decrease predicted risk and by how much on average. We interpret agreement between the two identification methods as increased confidence in stable signals, while discrepancies can indicate effects driven primarily by measurement availability/missingness, value gradients, or subgroup-specific contributions.

## Materials and methods

### Study cohort

This retrospective register-based study used routinely collected EHR data from Helsinki University Hospital (HUS), covering residents of the HUS catchment area from January 1, 2009 to November 7, 2024. The study cohort comprised (i) all infants admitted to a neonatal intensive care unit within HUS and (ii) infants whose mothers had documented use of predetermined medications during the nine months preceding birth. The medication lists are provided in S2 Table.

The initial dataset comprised 42,959 children. To ensure sufficient follow-up for outcome assessment, we restricted the cohort to children who were at least seven years old at the time of data extraction, yielding 21,118 children. As quality control, children with implausible gestational age values (*>* 46 weeks), missing maternal linkage information, missing birth weight, or invalid neuropsychiatric diagnosis dates were excluded, resulting in 21,087 children with valid birth and outcome information.

The initial 90 days of life served as the baseline for our predictions. After this period, a patient needed at least one recorded clinical encounter to be included in the cohort. Encounters encompassed any recorded diagnosis, laboratory measurement, hospital admission, or medication record. This approach targeted a subpopulation with observable healthcare utilization beyond the neonatal stage, resulting in a cohort of 18,332 children. The primary outcome was defined as the first major neuropsychiatric diagnosis occurring between 90 days and seven years of age. To maintain a consistent outcome assessment window and comparable follow-up across individuals, children whose first qualifying diagnosis was recorded only after age seven years were excluded. The final analytic cohort consisted of 17,655 children. An overview of the cohort selection process is shown in Fig 1.

**Fig 1.**
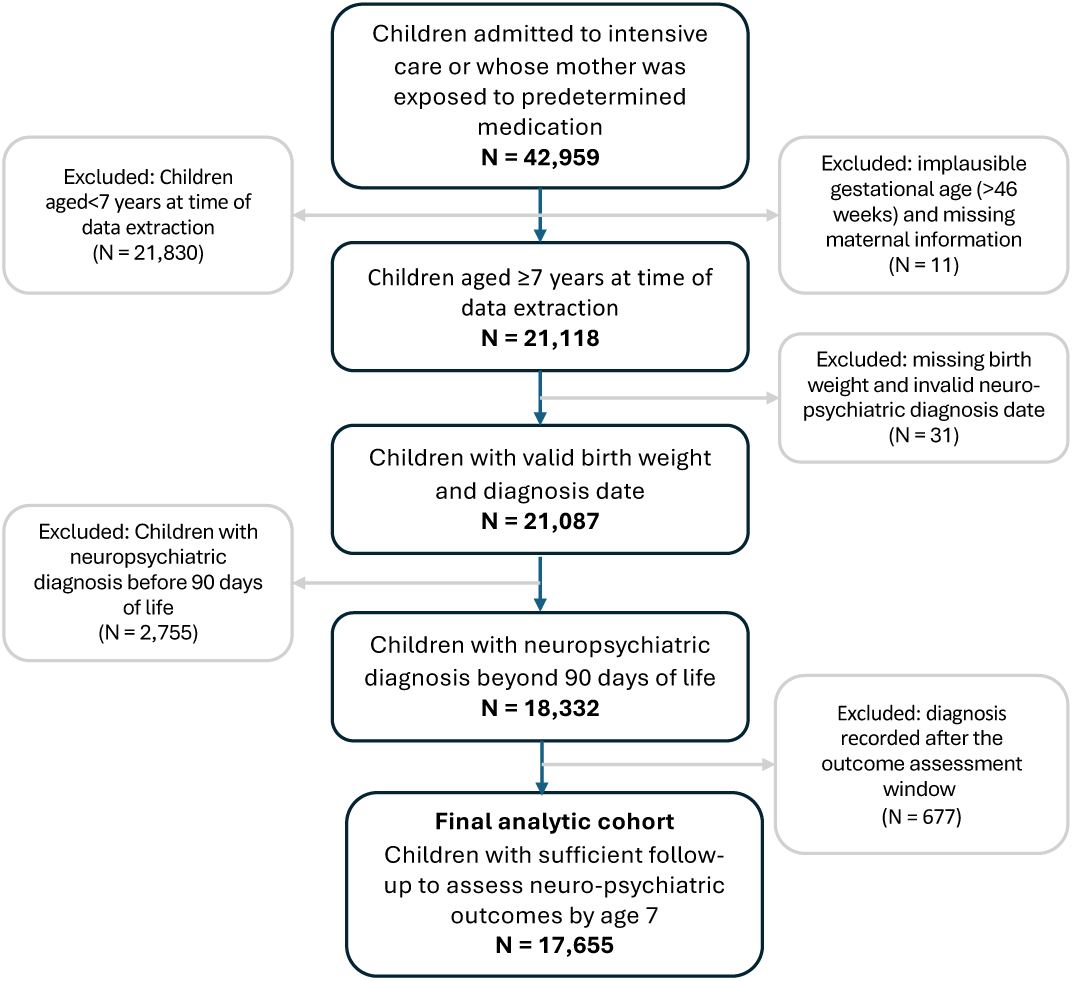
Cohort selection flow diagram. The diagram shows inclusion and exclusion steps used to derive the final analytic cohort (N=17,655) from the initial neonatal intensive care population.

All analyses were performed on a secure research platform at HUS (HUS Academic), complying with national data protection laws and the EU GDPR. The study was approved by HUS (§57/2024, §14/2025). According to national legislation, no further permissions or informed consents were needed for retrospective register-based study.

### Data preprocessing

We utilized EHR from neonatal intensive care and subsequent pediatric care including demographic and birth characteristics, maternal medication exposure during pregnancy, neonatal medication administration, clinical diagnoses, laboratory measurements, growth measurements, and treatment time in hospital, all linked at the individual child level (Table 1). Demographic and birth data included gender, date of birth, delivery method, gestational age, birth weight, birth length, head circumference, Apgar scores at 1, 5, and 10 minutes, number of fetuses, and body mass index (BMI). Maternal medication exposure during pregnancy was encoded as binary indicators reflecting exposure to medications of interest by monthly aggregation.

**Table 1.** Dataset variable summary for selected variables.

| Variable | Type | Undiagnosed<br>*N* (Obs.) | Diagnosed<br>*N* (Obs.) | Undiagnosed<br>Summary (SD) | Diagnosed<br>Summary (SD) | Adj.<br>*p*<sub>FDR</sub> |
| --- | --- | --- | --- | --- | --- | --- |
| **Demographics** |  |  |  |  |  |  |
| Head circumference at birth (cm) | Continuous | 13995 (13995) | 1125 (1125) | 34.20 (2.53) | 33.83 (2.81) | 1.59e-05* |
| Birth length (cm) | Continuous | 15442 (15442) | 1326 (1326) | 48.51 (4.19) | 47.26 (5.16) | 3.36e-23* |
| Birth weight (gr) | Continuous | 16235 (16235) | 1420 (1420) | 3131.53<br>(878.15) | 2898.84<br>(1014.14) | 7.01e-20* |
| Gestational age (days) | Continuous | 16235 (16235) | 1420 (1420) | 266.20<br>(23.37) | 260.39<br>(28.92) | 2.54e-17* |
| Apgar 10min | Ordinal (1-10) | 8175 (8175) | 745 (745) | 10.0 | 10.0 |  |
| Apgar 1min | Ordinal (1-10) | 16032 (16032) | 1400 (1400) | 9.0 | 9.0 |  |
| Apgar 5min | Ordinal (1-10) | 11441 (11441) | 1038 (1038) | 9.0 | 9.0 |  |
| Delivery Type | Categorical (1-4) | 16235 (16235) | 1420 (1420) | 1.0 | 1.0 |  |
| Gender | Categorical (1-2) | 16235 (16235) | 1420 (1420) | 2.0 | 2.0 |  |
| Number Of Fetuses | Ordinal (1-4) | 16235 (16235) | 1420 (1420) | 1.0 | 1.0 |  |
| **Growth** |  |  |  |  |  |  |
| BMI | Continuous | 15505 (31069) | 1336 (3875) | 12.78 (2.29) | 12.66 (2.27) | 0.007* |
| **Hospital** |  |  |  |  |  |  |
| Treatment time in hospital (days) | Presence | 16220 (158542) | 1420 (24550) | 5 | 6 |  |
| **Lab** |  |  |  |  |  |  |
| B-HKR (%) | Continuous | 14562 (94518) | 1313 (14408) | 43.51 (10.28) | 40.60 (10.04) | 3.70e-218* |
| B-Trom (E9/l) | Continuous | 14479 (87390) | 1308 (13817) | 288.87<br>(133.56) | 281.40<br>(144.49) | 1.31e-08* |
| E-MCHC (g/l) | Continuous | 14484 (87480) | 1308 (13824) | 347.75<br>(11.10) | 344.29<br>(11.56) | 2.41e-248* |
| E-MCV (fl) | Continuous | 14484 (87520) | 1308 (13835) | 98.38 (8.29) | 96.48 (8.81) | 1.03e-133* |
| E-Red cell distribution width (RDW) (%) | Continuous | 14171 (84337) | 1285 (13350) | 17.13 (2.66) | 17.64 (2.51) | 4.89e-97* |
| P-Ca-IonA (mmol/l) | Continuous | 13213 (98069) | 1221 (16545) | 1.32 (0.87) | 1.32 (0.20) | 0.847 |
| P-Krea (umol/l) | Continuous | 2234 (6957) | 421 (1687) | 56.44 (44.73) | 57.07 (43.27) | 0.788 |
| S-Fenob (umol/l) | Continuous | 121 (348) | 72 (322) | 103.02<br>(28.36) | 102.43<br>(29.20) | 0.923 |
| cB-pO2 (kPa) | Continuous | 12062 (72287) | 1139 (11752) | 6.50 (1.34) | 6.28 (1.44) | 1.38e-55* |
| uA-Be (mmol/l) | Continuous | 15163 (15163) | 1296 (1296) | -4.65 (3.85) | -4.58 (4.06) | 0.748 |
| uA-pO2 (kPa) | Continuous | 15162 (15162) | 1295 (1295) | 2.74 (1.17) | 2.74 (1.20) | 1.000 |
| Umbilical serum thyroid-stimulating hormone (uS-TSH) (mU/l) | Continuous | 16092 (16095) | 1396 (1396) | 8.39 (10.01) | 9.41 (29.62) | 0.017* |
| P-LupusAK (-) | Binary | 43 (44) | 32 (32) | 0.00 | 0.00 | 0.977 |
| **Child medications** |  |  |  |  |  |  |
| Phenobarbital (mg/kg/day) | Continuous | 126 (430) | 76 (401) | 9.08 (8.04) | 7.79 (7.23) | 0.052 |
| Levetiracetam (mg/kg/day) | Continuous | 33 (132) | 30 (274) | 21.44 (10.28) | 28.55 (15.08) | 9.27e-06* |
| **Diagnoses** |  |  |  |  |  |  |
| Other disturbances of cerebral status of newborn (P91) | Presence | 147 (168) | 75 (88) | 0.91% | 5.28% | 1.75e-43* |
| Congenital Malformations of Cardiac Septa (Q21) | Presence | 641 (871) | 128 (180) | 3.95% | 9.01% | 8.69e-18* |
| Congenital malformation syndromes (Q87) | Presence | 48 (51) | 30 (35) | 0.30% | 2.11% | 6.25e-21* |
| Chromosomal Abnormalities Not Elsewhere Classified (Q90) | Presence | 47 (70) | 58 (82) | 0.29% | 4.08% | 3.12e-68* |
| Encounters For Examinations Investigations (Z01) | Presence | 1608 (1753) | 313 (358) | 9.90% | 22.04% | 2.41e-43* |

*N* is the number of children with the variable observed, and Obs. is the total number of recorded occurrences across all children and time points. SD, standard deviation. Summary statistics are mean (continuous), median (categorical/ordinal; integer-coded), and percentage (presence). Adj. *p*<sub>FDR</sub> is the Benjamini-Hochberg false discovery rate (FDR)-adjusted p-value comparing undiagnosed and diagnosed groups; \* denotes Adj. *p*<sub>FDR</sub> < 0.05. An extended dataset variable summary is provided in S1 Table.

Medication data of neonates covered drugs administered during the early life, with particular focus on off-label medication use. Medication exposure was derived from time-stamped administration records and encoded longitudinally by daily cumulative dose per kg. The list of medications for mothers and children is given in S2 Table.

Diagnoses used for prediction were extracted from longitudinal EHR and included all diagnoses recorded during neonatal and subsequent pediatric care. Because routine clinical documentation may record the same underlying condition repeatedly across encounters, we consolidated diagnosis records temporally prior to modeling to reduce documentation-related redundancy. To harmonize diagnosis representation and reduce sparsity, International Classification of Diseases, 10th Revision (ICD-10) codes were truncated to the three-character category level. For example, Q63.1 and Q63.3 were grouped under Q63. We then applied a clinician-curated ICD-10 rule set to classify diagnosis categories as repeatable (e.g., acute events) versus non-repeatable or chronic; the ICD-10 blocks treated as repeatable and retained at monthly resolution are listed in S3 Table. For repeatable diagnoses, we retained longitudinal occurrences but merged repeated records of the same diagnosis within a one-month window from the first recorded instance into a single event; diagnoses recorded again after this window were retained as separate occurrences. For non-repeatable or chronic diagnoses, we retained only the first recorded occurrence and ignored subsequent records of the same diagnosis within the observation window.

Laboratory measurements consisted of time-stamped test results recorded during clinical care. These data were preprocessed to harmonize numeric formats, remove implausible values, and encode categorical or descriptive results.

Growth measurements, including weight and height, were extracted longitudinally and filtered using biologically plausible age-specific thresholds.

In addition, treatment time in hospital (days) was incorporated as a longitudinal signal. Each day a child was admitted to or present in the hospital was recorded as a binary presence indicator (1 = admitted/present), capturing duration and continuity of inpatient care over time. This representation reflects healthcare utilization intensity and complements clinical and medication data by encoding patterns of hospital exposure.

All data sources were aligned temporally and represented as individual-level longitudinal sequences covering the first 90 days of life. Although exact timestamps were available for most data sources, all observations were aggregated to daily resolution, as sub-daily variation may add noise for a prediction task targeting outcomes over a seven-year horizon. For non-sequential models that require fixed-length (static/vector) inputs, (i) neonatal medication exposure and treatment time in hospital were aggregated by summing total administered doses and total days of inpatient stay, respectively; (ii) laboratory measurements were summarized using predefined aggregation strategy, where selected tests were represented by clinically relevant extremes (maximum for S -Fenob, P-Krea, cB-Bil, P-AFOS, cB-pCO_2_ and P-AntiFXa, and minimum for cB-pH, and cB-HCO_3_-St), while all other laboratory variables were represented by the most recent observation within the observation window; and (iii) clinical diagnoses, growth measurements, and maternal medication exposure during pregnancy were represented using the most recent observation available within the observation window. This approach yields a fixed-length representation while retaining clinically relevant information.

The EHR feature set included 507 input variables, consisting of 497 time-dependent longitudinal variables—351 laboratory measurements, 115 diagnoses recorded during the neonatal period as covariates (excluding outcome-defining diagnoses), 17 child medications, 10 maternal medications during pregnancy, three growth measurements, and treatment time in hospital—and ten time-fixed demographic covariates.

Longitudinal inputs were constructed from time-stamped records, whereas demographic covariates were included as static features.

### Outcome definition

The primary outcome was the occurrence of *major neuropsychiatric morbidity* during follow-up. Major neuropsychiatric morbidity was defined based on a predefined set of neuropsychiatric diagnoses, listed in Table 2, identified from longitudinal clinical diagnosis records using ICD-10 codes aggregated at the three-character category level.

**Table 2.** Major neuropsychiatric morbidity conditions and ICD-10 codes.

| Condition | ICD-10 Code(s) |
| --- | --- |
| Cerebral palsy | G80.0–G80.9 |
| Epilepsy | G40.0–G40.9 |
| Intellectual disability | F70.0–F73.9, F78.0–F79.9 |
| Autism spectrum disorder | F84.0–F84.9 |
| Visual impairment | H53.0–H53.4, H54.0–H54.7 |
| Hearing impairment | H90.0–H90.8 |

For children with multiple neuropsychiatric diagnoses recorded during the follow-up, only the *first recorded diagnosis* was used to define the outcome event.

### Data analysis

This subsection describes the model development, evaluation, and interpretability procedures used in the study.

#### Model training and evaluation protocol

Unless otherwise noted, all predictive and interpretability results were obtained under a five-fold cross-validation protocol. In each fold, 20% of children formed the test set, and each child appeared in the test set exactly once across the five folds. After defining the test set for a fold, the remaining 80% of children were split once into 60% training and 20% validation, and this training/validation split was kept fixed for all model variants and analyses within that fold. The validation split was selected to match the composite outcome prevalence of the corresponding test split and was used for hyperparameter selection and early stopping. We performed three independent training runs per fold and selected the run with median validation area under the precision–recall curve (AUPRC) to obtain a robust fold-level estimate; Logistic Regression was trained once per fold due to its deterministic optimization. To compute stable global summaries (e.g., cohort-level performance curves and interpretability rankings), we concatenated test-set predictions across folds, yielding exactly one out-of-sample prediction per child.

#### Predictive modeling and benchmarking

The prediction task was formulated as a binary classification problem: predicting whether a child would receive any major neuropsychiatric diagnosis by age seven using longitudinal EHR data from the first 90 days of life.

Our primary model was STraTS, a time-aware Transformer-based framework designed for sparse and irregularly sampled clinical event data [15]. STraTS utilizes each observation as a triplet (*t, r, v*) corresponding to time, variable identity, and value, rather than a dense time*×*variable matrix. This event-based representation avoids time discretization and reduces reliance on imputation in the presence of high sparsity and irregular sampling [15]. Observation triplets are embedded using learnable variable embeddings together with continuous time–value embeddings, contextualized via multi-head self-attention, and aggregated through attention pooling into a patient-level representation for classification [15]. Following the original STraTS training protocol, the model was first pretrained using a self-supervised forecasting objective on the same cohort’s longitudinal data, where the model learns to predict masked or future observation values from surrounding context. The pretrained weights were then used to initialize the model for supervised fine-tuning on the binary classification task.

To benchmark STraTS against commonly used non-sequential models for structured clinical data, we evaluated Logistic Regression [25], Random Forest [23], and eXtreme Gradient Boosting (XGBoost) [26]. Because these baseline models require fixed-length feature representations, longitudinal records from the first 90 days of life were converted into static summary features, as described in the Data Preprocessing subsection.

Hyperparameters for all models were tuned using random search over predefined search spaces, with configurations selected based on validation performance. For the baseline models, we tuned standard regularization and tree-based parameters and addressed class imbalance using class weighting where applicable. Both mean and median imputation strategies were evaluated during baseline development, and the method yielding the best validation performance was selected separately for each baseline model. To ensure a fair comparison, the same child-level training, validation, and test splitting protocol was applied across all models to prevent information leakage across individuals. Full hyperparameter search spaces and optimization details are provided in S4 Table.

Model performance was evaluated using AUPRC and area under the receiver operating characteristic curve (AUROC). Because the outcome is highly imbalanced, we treat AUPRC as the primary metric, as it more directly reflects performance on the positive class; AUROC is reported as a complementary, threshold-independent summary of ranking performance. In imbalanced settings, AUROC can remain high even when precision is low, whereas AUPRC is sensitive to false positives and therefore better captures clinically relevant positive-prediction performance.

#### Model interpretability and feature effect analysis

The original STraTS framework includes an interpretable variant (iSTraTS) [15] that provides instance-level attention-based explanations by highlighting which input observations most influence each individual prediction. However, iSTraTS does not directly support cohort-level variable importance rankings or systematic analysis of how variable values relate to predicted risk across the population. To address these complementary interpretability needs, we conducted post hoc interpretability analyses for STraTS using three approaches that independently characterize model prediction at both the individual and cohort level: perturbation-based variable importance, LOO feature attribution, and value-dependent effect analysis.

Notably, the triplet-based input representation described above naturally supports variable ablation: removing a variable’s observations simply shortens the input sequence, avoiding masking or imputation artifacts. This property facilitates both the perturbation and LOO analyses described below.

### Perturbation-based variable importance

Perturbation-based variable importance was quantified using *feature permutation* [23, 27] and *ablation* [28], and importance was defined as the resulting decrease in predictive performance. For time-fixed demographic covariates, permutation importance was applied by randomly permuting each variable across children and recomputing performance, thereby breaking its association with the outcome while preserving its marginal distribution. For all time-dependent variables (medications, diagnoses, laboratory and growth measurements, and treatment time in hospital), ablation-based importance was applied by removing a variable from the model input and evaluating the corresponding change in test-set performance.

The ablation analysis was conducted at two levels of granularity. First, ablation was applied one variable at a time to obtain an importance ranking over individual features. Second, a source-level analysis was performed by ablating all variables belonging to a given data source (e.g., all laboratory measurements or all diagnoses) to quantify the relative contribution of each modality. Finally, because in our setting phenobarbital medication exposure and measured phenobarbital serum concentration capture correlated information from medication records and laboratory measurements, an additional grouped ablation removed both features simultaneously to assess their combined contribution.

### Value-dependent variable effects

To quantify how different values or levels of individual variables influence model predictions at a global level, we performed a value-dependent variable effect analysis. The analysis is conceptually related to partial dependence analysis [24], but is implemented in a model-agnostic manner suitable for complex nonlinear models, and it is intended to characterize model reliance on variable values.

For continuous variables, model predictions were evaluated at five quantile levels (Q10, Q30, Q50, Q70, Q90) while keeping all other covariates unchanged. For time-dependent continuous variables, all observed values of that variable within each child’s trajectory were set to the same quantile level for each evaluation, while preserving all other variables. The reported effect is computed as the prediction at Q90 minus the prediction at Q10, so that a positive value indicates that higher variable values increase predicted risk. For ordinal variables, predictions were compared between lower and higher levels; for nominal categorical variables, we used pre-defined category contrasts (gender: 1:female, 2:male; birth type: 1:vaginal vs 2–4 assisted/operative delivery types), and for presence variables, effects were assessed using an ablation contrast. For a presence variable *j*, the individual-level effect was defined as

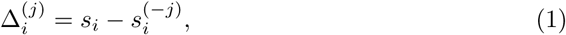

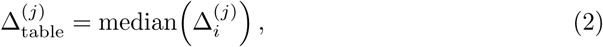

where *s_i_ ≡ f(x_i_)* is the model output (on the logit scale) for child i under the original input x_i_, and *s_i_^(-j)^ ≡ f(x_i_^\j^)* is the output after ablating variable j. Under this convention, positive Δ*_i_*^(*j*)^ indicates that the presence of variable *j* increases the predicted risk for child *i*.

To summarize stability in the direction of the effect across children, we report *Consistency*,

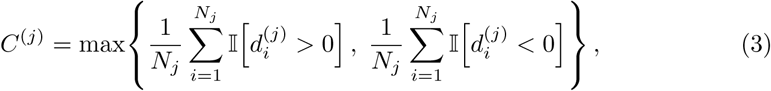

where *N_j_* is the number of children with observations of variable *j*, I[*·*] is the indicator function, and d*_i_*^(*j*)^ captures the direction of the effect for child *i*. For presence variable *j*, *d_i_*^(*j*):^ Δ*_i_*^(*j*)^. For continuous, ordinal, and categorical variables, d*_i_*^(*j*)^ is the per-child linear slope fitted to the logit response over the evaluated levels:

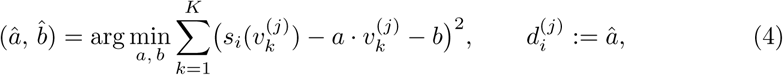

where *s_i_*(*v_k_*^(*j*)^) is the model output on the logit scale for child *i* when variable *j* is set to *v_k_*^(*j*)^ while all other variables have their original values. For continuous variables, *v_k_*^(*j*)^ corresponds to quantile levels (Q10, Q30, Q50, Q70, Q90; *K* = 5); for ordinal variables, *v_k_*^(*j*)^ corresponds to the score levels; for categorical variables, *v_k_*^(*j*)^ corresponds to the pre-defined category contrasts described above, so a positive *d_i_*^(*j*)^ indicates that the higher coded category increases predicted risk.

Variable effects are reported on two complementary scales: changes in the model output on the logit scale and changes in predicted risk (probability) of outcome. We summarize each effect using the median change and interquartile range (IQR), providing a robust measure of both magnitude and variability. The fraction of perturbations with consistent directional effects is additionally reported to indicate stability across children.

### Leave-one-out feature attribution

To characterize which inputs most strongly influenced individual model predictions, we performed a post hoc interpretability analysis using LOO feature attribution [28, 29], also known as occlusion-based attribution. While perturbation-based importance measures how much a variable contributes to overall model discrimination (via cohort-level performance change), LOO attribution measures how much a variable shifts the prediction for each individual patient, providing both local (instance-level) and global (cohort-level) interpretation.

For each individual *i* and feature *j*, we computed the LOO attribution Δ^(*j*)^ as defined in Eq. 1. For time-dependent variables, removal was implemented by dropping all triplets of that variable from the individual’s input sequence, leveraging STraTS’s ability to process varying-length sequences; for demographic features, the feature value was masked. Positive values indicate that the feature increases the model output for that individual, whereas negative values indicate a decrease. We ranked features by their mean absolute attribution,

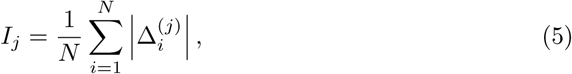

where *N* denotes the number of evaluated individuals.

Unlike Shapley-value-based methods [30], which average marginal contributions over all possible feature subsets [28], LOO attribution evaluates each feature’s contribution only in the context of all other features being present. This makes LOO attribution computationally tractable for high-dimensional inputs (507 features in our setting), but means that for correlated features, LOO attribution may underestimate the importance of redundant features because the model can compensate through correlated inputs that remain present.

Because global importance *I_j_*averages *|*Δ^(*j*)^*|* over all individuals, it reflects both (i) the per-individual effect size when the feature is informative and (ii) how frequently the feature is present or varies in the cohort. In high-dimensional and sparse representations (common in EHR-derived data), rare exposures may therefore appear less prominent in global LOO rankings despite potentially large effects among the subset of individuals for whom they occur. We therefore interpret global LOO rankings as cohort-level summaries and complement them with perturbation-based importance, which measures the effect of each variable on overall model discrimination regardless of its prevalence in the cohort.

### Visualization of patient trajectory representations

STraTS generates multiple representation levels, including event-level contextual embeddings, a pooled patient-level time-series embedding, and a separate demographic embedding that is concatenated with the time-series embedding for final prediction. For visualization, we used only the pooled 8-dimensional patient-level time-series embedding (derived from time-varying inputs), hereafter referred to as the *event sequence representation*, to assess whether the learned latent representation captures clinically plausible patient trajectories. We projected this representation into two dimensions using Principal Component Analysis (PCA) to examine how perinatal characteristics relate to global structure in the representation space.

## Results

### Data summary

The final cohort comprised 17,655 children, of whom 8.0% (1,420/17,655) received a major neuropsychiatric diagnosis by age seven. Among the 1,420 diagnosed children, classified by their earliest recorded diagnosis, autism spectrum disorder was the most common subtype (430; 30.3%), followed by visual impairment (336; 23.7%), hearing impairment (203; 14.3%), intellectual disability (173; 12.2%), epilepsy (163; 11.5%), and cerebral palsy (115; 8.1%). Children diagnosed with major neuropsychiatric conditions had lower mean birth weight (2,899 vs. 3,132 g), shorter mean gestational age (260 vs. 266 days), and shorter mean birth length (47.3 vs. 48.5 cm) compared to undiagnosed children (Table 1). Median Apgar scores at 1 and 5 minutes were 9 in both groups.

Selected neonatal diagnoses were substantially more prevalent among diagnosed children, including chromosomal abnormalities (Q90; 4.1% vs. 0.3%) and other disturbances of cerebral status of newborn (P91; 5.3% vs. 0.9%).

### Predictive modeling and benchmarking

Model performance on the held-out test folds is summarized in Table 3. Given the class imbalance, we report AUPRC as the primary metric and AUROC for completeness [31]. For reference, the expected AUPRC of a non-informative (random) classifier equals the positive-class prevalence, i.e., 8.0% in this cohort. Across models, STraTS achieved the highest AUPRC, indicating improved identification of children at risk compared to baseline methods. Among the baseline models, Random Forest showed the strongest overall performance, outperforming Logistic Regression and XGBoost in terms of AUPRC.

**Table 3.** Benchmark model comparison on the test set.

| Model | Test AUPRC (mean ± SD) | Test AUROC (mean ± SD) |
| --- | --- | --- |
| Logistic Regression | 0.151 ± 0.007 | 0.615 ± 0.011 |
| Random Forest | 0.166 ± 0.008 | 0.630 ± 0.011 |
| XGBoost | 0.128 ± 0.010 | 0.592 ± 0.010 |
| STraTS | 0.171 ± 0.022 | 0.607 ± 0.017 |

Performance is reported as mean ± SD across cross-validation folds. Due to class imbalance, AUPRC is emphasized as the primary evaluation metric.

Notably, STraTS achieved the highest AUPRC (0.171) but a lower AUROC (0.607) than both Random Forest (0.630) and Logistic Regression (0.615). This divergence is consistent with the known behavior of these metrics under class imbalance: AUPRC is more sensitive to improvements in minority-class ranking, whereas AUROC reflects discrimination across both classes and can be dominated by the large number of majority-class comparisons [31]. The AUPRC difference between STraTS and Random Forest was small in absolute terms (0.171 vs. 0.166), corresponding to an approximately 3% relative improvement, while STraTS showed a larger gain over Logistic Regression (approximately 13%; 0.171 vs. 0.151). STraTS also exhibited higher variability across folds (AUPRC SD; 0.022) compared to the baseline models (0.007–0.010), likely reflecting the greater sensitivity of transformer-based models to fold composition given the small number of positive cases per fold, i.e. approximately 284 per test fold.

**Fig 2.**
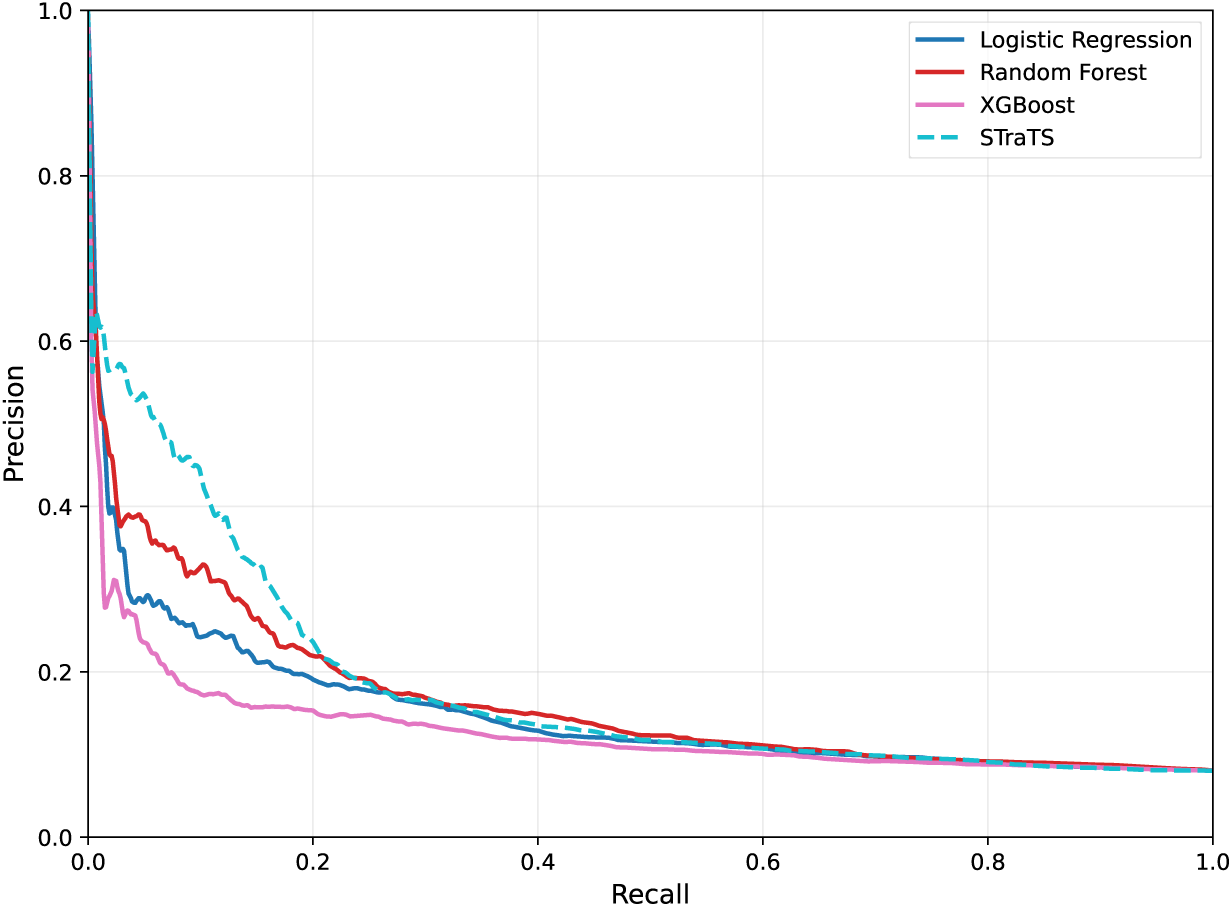
Mean precision–recall curves on the test set. Curves show mean precision–recall performance across cross-validation folds for Logistic Regression, Random Forest, XGBoost, and STraTS. Precision–recall analysis is emphasized due to the imbalanced outcome distribution.

Fig 2 illustrates mean precision–recall performance on the held-out test folds, highlighting improved minority-class discrimination for STraTS compared to baseline models. The mean receiver operating characteristic (ROC) curve, as well as precision–recall and ROC curves with cross-fold variability bands, are provided in S1 Fig.

### Model interpretability and feature effect analysis

#### Perturbation-based variable importance

Fig 3a summarizes global importance as the relative change in test-set AUPRC. At the source level, removing all laboratory variables or all diagnosis variables led to the largest performance decreases (approximately *−*20% relative to baseline AUPRC), indicating that these two modalities account for the dominant share of discriminative signal in the model. Ablating treatment time in hospital resulted in a smaller but non-negligible drop, suggesting that care-context features contribute additional predictive information beyond clinical measurements and diagnoses.

Among time-fixed covariates, *birth weight* and *gender* exhibited the strongest importance, each producing a clear reduction in AUPRC when disrupted, while *APGAR score at 1 minute* showed a more modest but consistent effect. Within the longitudinal variables, the single largest performance decrease was observed for *chromosomal abnormalities (Q90)*, consistent with this diagnosis acting as a strong marker of later neuropsychiatric risk. Other neonatal-condition diagnoses (e.g., other disturbances of cerebral status of newborn-*P91* and congenital malformations of cardiac Septa-*Q21*) also showed measurable importance, although with smaller effect sizes. Z01 (encounters for examinations/investigations) also showed a strong global importance, consistent with care-process/utilization patterns contributing to discrimination.

Medication-related features displayed moderate global effects overall; however, *phenobarbital medication* related variables were consistently among the most influential medication-associated signals. In particular, the grouped phenobarbital ablation (combining medication and laboratory representations) produced a larger performance decrease than either representation alone, suggesting that phenobarbital-associated information is distributed across correlated inputs. In contrast, several laboratory and medication variables (e.g., red cell distribution width (RDW), umbilical arterial blood base excess (Ua-be), levetiracetam) had effects close to zero, indicating limited contribution to overall discrimination at the cohort level.

#### Value-dependent variable effects

Building on the perturbation-based variable effect analysis (Fig 3a), we examined value-dependent effects for a selected subset of variables that showed the largest and most stable performance impact when removed or permuted. From the full set of variables, we focused on these variables to translate global importance into clinically interpretable effect magnitudes, quantifying how changes in a variable’s value (or presence) shift the model’s predicted neuropsychiatric risk (Fig 3b, Table 4).

**Table 4.** Value-dependent effects of selected variables (Fig. 3a).

| Variable | N | Consist. | $\Delta$ logit | $\Delta$ risk in pp | OR |
| --- | --- | --- | --- | --- | --- |
| **Categorical/Ordinal/Binary variables** |  |  |  |  |  |
| *Gender*<sup>†</sup> | 17,655 | 97% pos | 0.48 (0.32) | 0.77 (0.68) | 1.62 |
| *APGAR (1 min)*<sup>†</sup> | 17,655 | 76% neg | -0.24 (0.44) | -0.44 (1.05) | 0.79 |
| Lupus antibody | 74 | 85% pos | 2.12 (4.23) | 1.88 (16.9) | 8.30 |
| **Continuous variables** |  |  |  |  |  |
| *Birth weight*<sup>†</sup> | 17,655 | 98% neg | -0.82 (0.58) | -1.56 (1.69) | 0.44 |
| Phenobarbital | 202 | 77% pos | 0.01 (0.04) | 0.04 (0.21) | 1.01 |
| Phenobarbital (serum) | 193 | 92% pos | 0.21 (0.63) | 0.94 (4.88) | 1.24 |
| Creatinine | 2,637 | 73% neg | -0.01 (0.02) | -0.02 (0.06) | 0.99 |
| uS-TSH | 17,469 | 66% pos | -0.00 (0.02) | -0.01 (0.05) | 1.00 |
| **Presence variables** |  |  |  |  |  |
| Chromosomal abnormalities (Q90) | 103 | 100% inc | 5.57 (3.94) | 82.9 (36.7) | 263 |
| Other disturbances of cerebral status of newborn (P91) | 217 | 99% inc | 1.16 (1.48) | 6.02 (20.1) | 3.18 |
| Encounters for exams or investigations (Z01) | 1,569 | 99% inc | 0.13 (0.15) | 0.28 (0.53) | 1.14 |
| Congenital Malformations of Cardiac Septa (Q21) | 718 | 90% inc | 0.21 (0.46) | 0.32 (0.96) | 1.23 |
| Treatment time in hospital (days) | 17,622 | 77% inc | 0.05 (0.12) | 0.07 (0.23) | 1.05 |

N: children with the feature observed. Consist.: fraction of children with the dominant effect direction, $\max\{\Pr(\Delta_i > 0), \Pr(\Delta_i < 0)\}$ . Effects are computed as high minus low (Q90 minus Q10 for continuous, higher minus lower category for categorical) or present minus absent (for presence variables); positive $\Delta$ indicates increased predicted risk. $\Delta$ logit: change on the logit scale; $\Delta$ risk in pp: change in predicted risk in percentage points; both reported as median (IQR) across children. OR: odds ratio corresponding to $\Delta$ logit. <sup>†</sup>Demographic (time-fixed) variable.

Diagnostic indicators reflecting early-life severity and complex neonatal conditions show the strongest risk-increasing effects. Chromosomal abnormalities (Q90) exhibit the largest shift in predicted risk when present (median Δ risk +82.9 percentage points; odds ratio [OR] 263), and other disturbances of cerebral status of newborn (P91) also show a substantial increase (median Δ risk +6.02 percentage points; OR 3.18).

Congenital malformations of cardiac septa (Q21) and encounters for examinations and investigations (Z01) demonstrate smaller but consistent risk-increasing effects (Table 4).

Demographic and early physiological variables exhibit more moderate but highly consistent associations. Higher birth weight is associated with reduced predicted risk (median Δ risk *−*1.56 percentage points; OR 0.44; 98% negative direction), and higher Apgar score at 1 minute shows a protective association (median Δ risk *−*0.44 percentage points; OR 0.79; 76% negative direction). Gender shows a consistent higher risk (positive direction) towards male gender (Table 4).

Medication-related variables showed smaller effects on the probability scale but can display consistent directionality in specific subgroups. Phenobarbital exposure and serum concentration measurements were overall associated with increased risk, although the median effect for exposure to medication alone is close to zero (Table 4).

Laboratory measurements such as uS-TSH showed near-zero effects on predicted risk at the population level, despite moderate directional consistency, suggesting limited contribution to global risk estimates within the model (Table 4).

**Fig 3.**
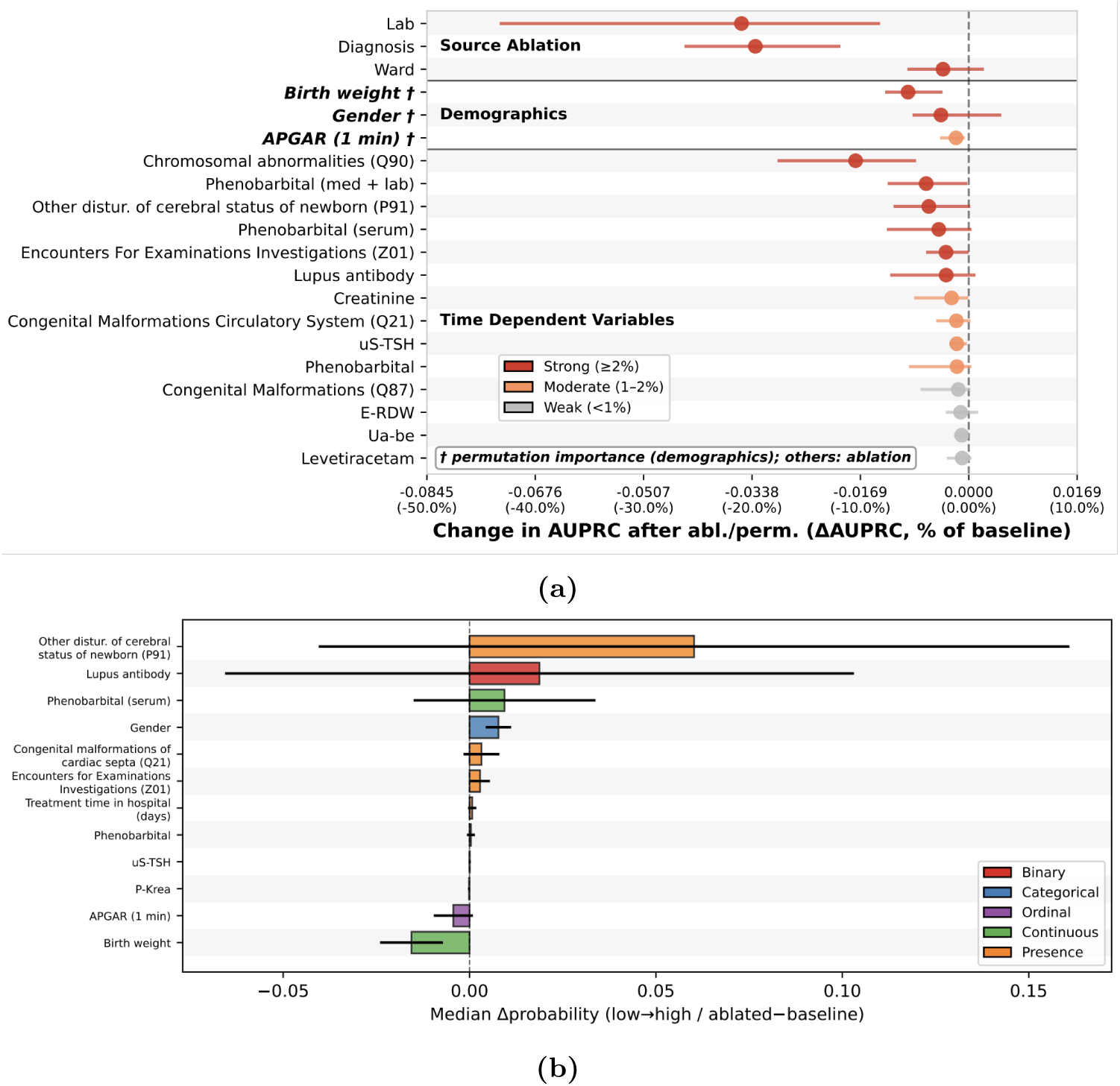
Variable importance and value-dependent effects. **(a)** Permutation-based variable importance. Mean relative change in test-set AUPRC after removing or permuting each variable (averaged across folds); horizontal bars show variability. More negative values indicate higher importance; dashed line denotes no change. **(b)** Value-dependent effects. Median change in predicted probability (Δ*p*) for selected variables (Table 4). Effects compare Q10 vs Q90 for continuous variables, lower vs higher category for categorical, and presence vs absence for presence variables. Points: median; bars: IQR. Positive values indicate increased predicted risk, e.g. in case of gender, males have higher risk (female= 1, male= 2).

Overall, the value-dependent variable analysis complements the perturbation results by translating variable importance into effect magnitudes that are easier to interpret clinically. The two-scale visualization in Fig 3b facilitates simultaneous inspection of dominant and near-zero effects, supporting transparent interpretation of model prediction.

#### Leave-one-out feature attribution

Fig 4a summarizes LOO attribution-based global interpretability for the combined feature set (demographics and time-dependent variables). Variables identified as globally important by LOO attribution show *partial* overlap with those highlighted by the perturbation-based importance analysis; specifically, five variables were independently identified by both perturbation and LOO attribution, and all five were further confirmed through value-dependent effect characterization (S5 Table). This agreement between the two independent identification methods indicates that the core set of influential features is stable with respect to the choice of explanation technique.

**Fig 4.**
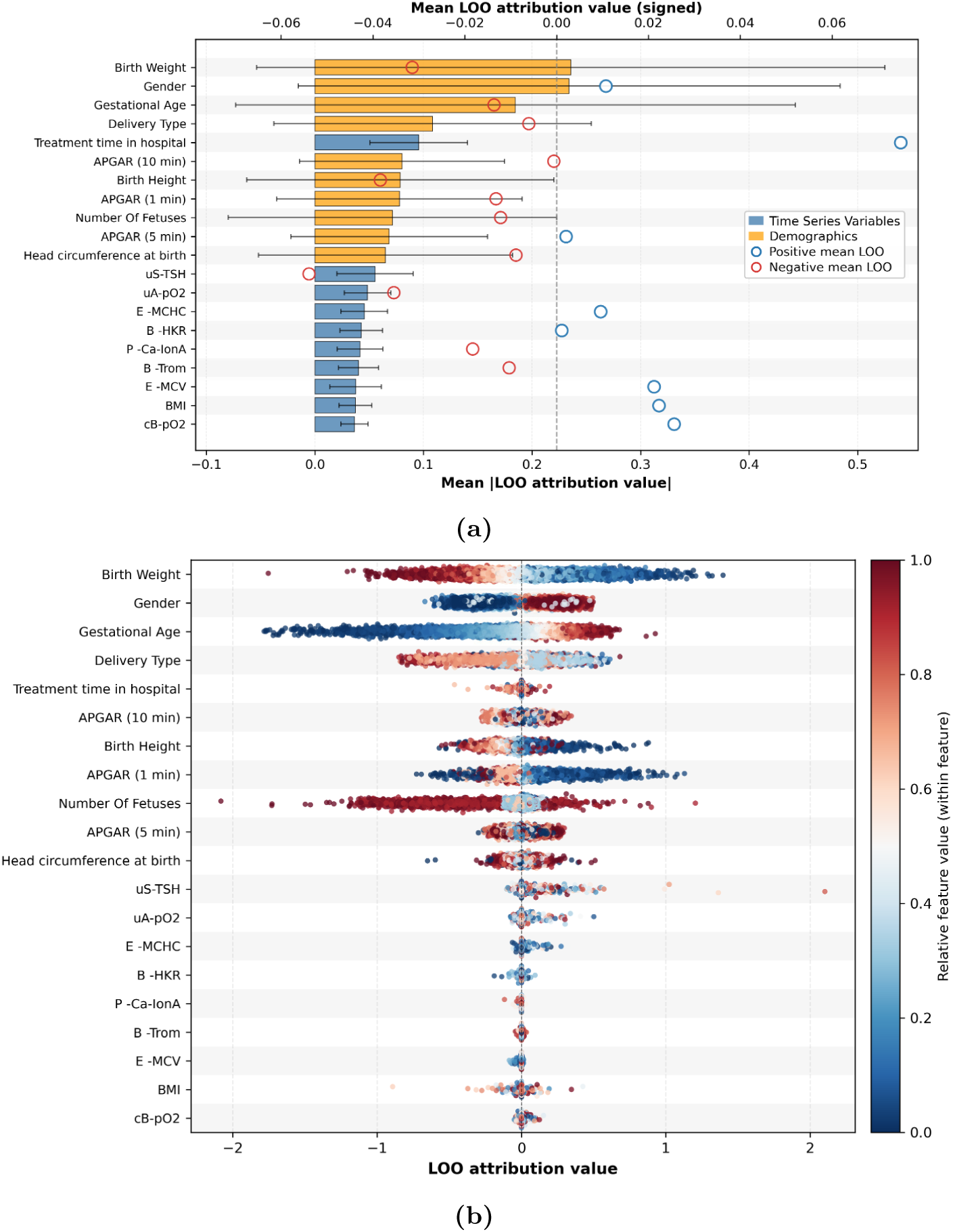
LOO attribution-based interpretability for the top 20 variables (demographics + time-dependent). (a) Global LOO importance with direction. Bars show mean absolute LOO attribution values for the positive class, averaged over the test set; variables are ranked by mean *|*Δ*|*. Circles indicate the mean signed attribution value, summarizing average direction, and whiskers indicate variability across individuals. (b) LOO attribution summary (beeswarm) plot for the same variables. Each point represents one child in the test set. The x-axis shows the LOO attribution value for the positive class; positive values increase and negative values decrease the risk for that individual. Point color indicates the feature value scaled within each variable (e.g., within-feature percentile; low to high); for categorical variables, color reflects the encoded category.

Among time-dependent variables, features related to care context (e.g., treatment time in hospital) and selected laboratory and growth measurements appear among the highest-ranked contributors in the global LOO attribution summary. The beeswarm visualization (Fig 4b) illustrates the distribution of feature attributions across individuals and highlights substantial heterogeneity in per-child contributions underlying these global summaries.

Directionality patterns in the LOO attribution plots show that the same variable may contribute positively or negatively to the model output across different individuals, depending on feature value and the broader input context. For categorical variables (e.g., gender and delivery type), LOO attributions exhibit clustered patterns corresponding to encoded category values. As noted in the Methods, global LOO importance averages attributions across all individuals, which can underweight rare features; we discuss the implications for specific variables in the Discussion.

Notably, *gestational age* exhibits a reversed directionality in the LOO attribution beeswarm plot (Fig 4b): higher values are associated with positive LOO contributions (increased predicted risk), contrary to the established clinical relationship and the cohort-level association in our data (mean 260 vs. 266 days in diagnosed vs. undiagnosed children; Table 1). We attribute this to a feature redundancy artifact and discuss it in the Discussion.

#### Visualization of patient trajectory representations

Fig 5 presents a PCA projection of the event sequence representations learned using only time-varying variables on the test set. Children diagnosed with major neuropsychiatric disorders show increased density in specific regions of the embedding space (Fig 5 A), which closely aligns with areas of elevated predicted risk from the STraTS model (Fig 5 B), indicating consistency between the learned representation and model predictions.

Overlays of time-dependent clinical characteristics reveal additional structure.

Regions associated with longer sequence length (i.e., a higher number of time-stamped variable observations) and greater total time spent in hospital largely overlap and coincide with higher predicted risk (Fig 5 C–D). In contrast, overlays of birth weight and gestational age show an opposite pattern, with lower values aligning with regions of elevated risk (Fig 5 E–F), consistent with established perinatal risk factors.

**Fig 5.**
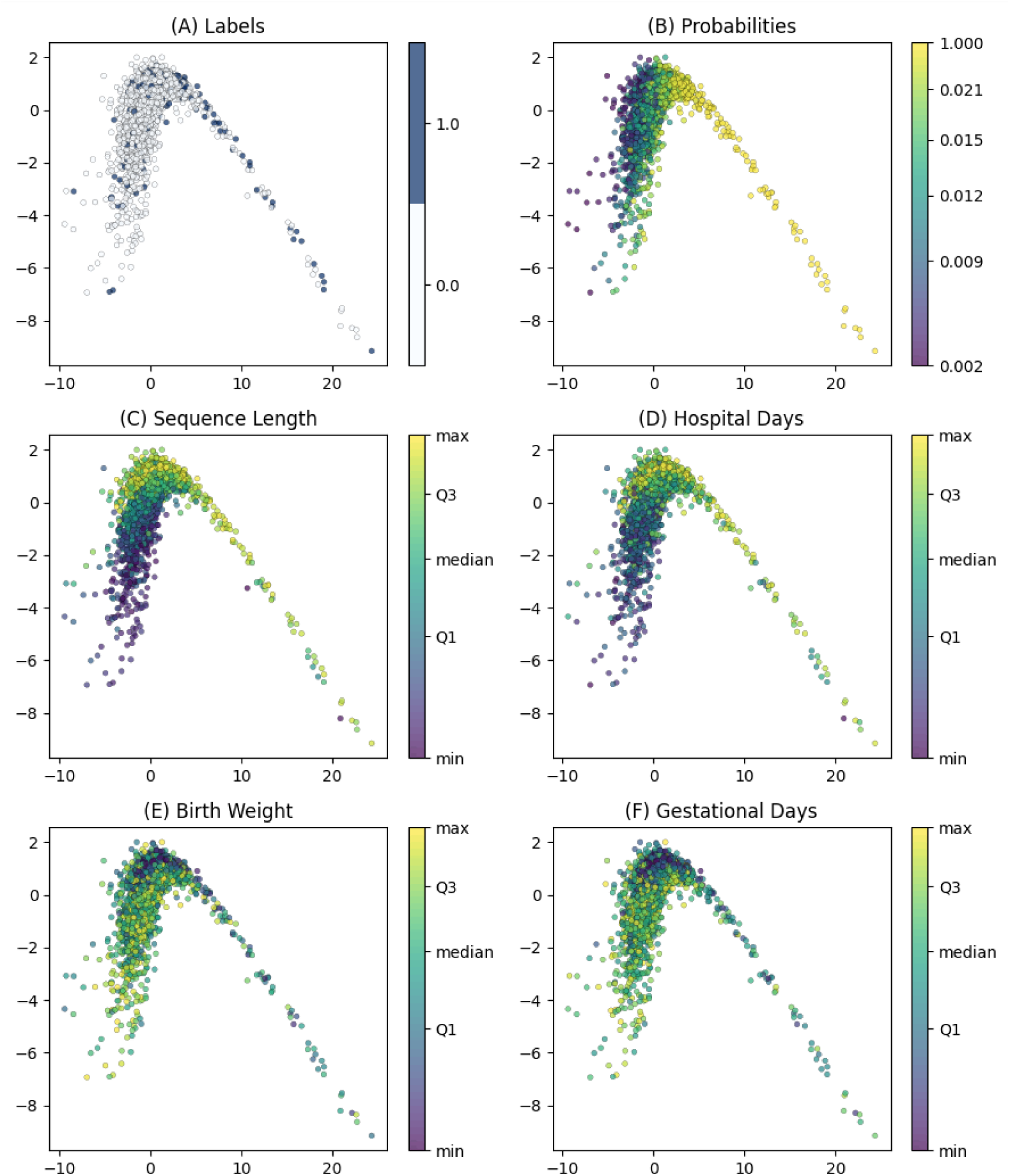
PCA projection of the eight-dimensional event sequence representations (test set). Each point represents a child. The top panels show (A) the true outcome label (major neuropsychiatric diagnosis vs. none) and (B) the predicted risk from the STraTS model. The remaining panels overlay selected clinical and perinatal characteristics, including (C) sequence length, (D) total hospital days, (E) birth weight, and (F) gestational age. Point color encodes the feature value scaled by its empirical distribution.

## Discussion

In this study, we applied a time-aware transformer model (STraTS) to longitudinal neonatal EHR data from the first 90 days of life to predict major neuropsychiatric diagnoses by age seven in a cohort of 17,655 at-risk children (Fig 1). Our objectives were to evaluate the predictive performance of sequence-based modeling on this task and to assess whether complementary explanation approaches can yield clinically meaningful and stable interpretability findings. The results demonstrate that (1) STraTS achieved the highest AUPRC among all evaluated models, indicating improved minority-class discrimination; (2) two independent variable identification approaches—perturbation-based importance and LOO feature attribution—converged on a core set of clinically plausible predictors, and value-dependent effect analysis further characterized the direction and magnitude of the perturbation-selected variables; and (3) the learned event sequence representations captured clinically coherent patient structure aligned with established perinatal risk dimensions.

The neuropsychiatric outcomes examined in this study—cerebral palsy, epilepsy, intellectual disability, autism spectrum disorder, and sensorineural impairment—have multifactorial etiologies involving combinations of genetic susceptibility [32–34], prenatal exposures including maternal medications and illnesses, complications during delivery, neonatal morbidity, and later environmental factors such as infections, trauma, and psychosocial adversity [2, 5, 11]. Despite the multifactorial nature of these outcomes, our model outperformed all baseline classifiers using only EHR data from the first 90 days of life, demonstrating that neonatal clinical trajectories contain identifiable and actionable signals for long-term neuropsychiatric risk. These results indicate that sequence-based modeling of routinely collected neonatal data can identify infants at elevated risk based on neonatal-period exposures, providing a foundation for early risk stratification in clinical practice.

STraTS achieved the highest AUPRC (0.171; Table 3), improving over Random Forest (0.166), while all models substantially outperformed the non-informative baseline (AUPRC = 0.08). The absolute gain over Random Forest reflects the inherent difficulty of the prediction task: the outcome is rare (8% prevalence), etiologically heterogeneous, and determined in large part by factors not observable in neonatal EHR data.

Nevertheless, AUPRC is the metric most sensitive to minority-class ranking under class imbalance [31], and the consistent advantage of STraTS on this metric indicates that sequence-based modeling of irregularly sampled longitudinal data provides incremental discriminative value beyond static summary representations, despite higher fold-to-fold variability (Table 3).

Interpretability is crucial for evaluating model performance in the medical domain and for gaining meaningful insights from data. The three complementary interpretability methods (described in detail in the Methods section) converged. Five variables were independently identified by both perturbation-based importance—which provides a cohort-level ranking insensitive to how the model internally distributes signal among correlated inputs—and LOO attribution. All five were further confirmed through value-dependent effect characterization (S5 Table), indicating that the core set of influential predictors is stable with respect to the choice of identification technique. However, these rankings characterize model reliance on variables associated with neuropsychiatric risk in aggregate, because the composite outcome groups etiologically distinct conditions with substantial within-condition heterogeneity (discussed below). Variables with broad cross-condition relevance—such as birth weight, male sex, and chromosomal abnormalities—are likely overrepresented, whereas subtype-specific predictors may be diluted.

The identified variables are consistent with established clinical knowledge. The strongest risk-increasing effects were observed for severe early diagnoses: chromosomal abnormalities (Q90; OR 263) and neonatal cerebral-status disturbances (P91; OR 3.18) showed large and highly consistent effects, in line with their known associations with long-term neurodevelopmental impairment [2, 8, 35]. Among demographic variables, higher birth weight was associated with reduced risk (OR 0.44; 98% consistency), consistent with the well-established protective effect of appropriate fetal growth [3, 4, 36].

Male gender showed a consistent risk-increasing effect (OR 1.62; 97% consistency), aligning with the higher prevalence of several neuropsychiatric conditions in males [2, 37]. Healthcare utilization features—treatment time in hospital and encounter codes (Z01)—contributed to discrimination, likely reflecting the intensity and complexity of neonatal care trajectories rather than direct causal effects [38].

Phenobarbital-related variables (medication exposure and maximal serum phenobarbital concentrations) showed consistent risk-increasing directionality, particularly when assessed jointly; this signal likely indexes the severity of underlying neonatal seizure activity rather than a direct medication effect [8, 9].

Notably, global LOO attribution rankings underweighted rare but clinically severe conditions such as chromosomal abnormalities (Q90) and cerebral-status disturbances (P91), because mean absolute attribution values average over all individuals and thus dilute large per-individual effects of rare exposures. In contrast, perturbation-based ablation, which evaluates the model’s response to complete removal of a variable via cohort-level performance change, captured the strong predictive reliance on these conditions. This complementarity demonstrates that no single interpretability method provides a complete picture in high-dimensional, sparse EHR settings [39–41]. For rare but important clinical markers, perturbation-based approaches may be more informative at the cohort level, while LOO attribution remains valuable for characterizing heterogeneity and context dependence at the individual level.

The LOO attribution analysis assigned higher gestational age a risk-increasing direction, opposite to the established clinical relationship [2, 3] and the cohort-level association in our data (Table 1). We attribute this to a feature redundancy artifact inherent to LOO attribution. LOO evaluates each feature’s contribution in the context of all other features remaining present. Because birth weight is strongly correlated with gestational age, it already captures the protective prematurity-related signal when gestational age is removed. The residual LOO attribution for gestational age therefore reflects only its conditional contribution given that birth weight is present, which can have an unintuitive direction. Birth weight consistently showed the expected protective direction—identified by both perturbation and LOO attribution and confirmed by value-dependent effect characterization (98% consistency; Table 4). This discrepancy was detectable precisely because gestational age appeared only in the LOO attribution ranking and not in the perturbation-based analysis (S5 Table). Reliance on a single identification method could either miss such artifacts or present clinically implausible findings without diagnostic context.

The event sequence representation analysis provided further qualitative support for the clinical plausibility of the learned representations. Children with similar longitudinal observation profiles and perinatal characteristics were located closer to one another in the representation space, and regions of elevated predicted risk aligned with longer hospitalization, higher sequence length, lower birth weight, and lower gestational age—all established perinatal risk dimensions. This suggests that the learned event sequence representations capture clinically meaningful structure related to temporal exposure intensity and neuropsychiatric risk, beyond what is reflected in classification metrics alone.

While prior work has largely focused on near-term outcomes or specific conditions using static perinatal variables [20, 42–44], our study addresses this gap. The principal novelty lies not in applying a particular model or interpretability method, but in demonstrating *when and why* different explanation approaches diverge on the same clinical prediction task—and what those divergences reveal about model prediction in sparse, high-dimensional EHR data. Specifically, the comparison between perturbation-based importance and LOO attribution exposed two concrete failure modes that would have gone undetected under a single-method framework: the systematic underweighting of rare but clinically severe conditions in global LOO attribution rankings, and the feature-redundancy attribution artifact for gestational age. These are not theoretical concerns but practical pitfalls with direct implications for how interpretability findings are communicated to clinicians. The value of the multi-method framework is therefore diagnostic. It provides internal checks that distinguish robust signals from method-specific artifacts, supporting more trustworthy interpretation of model prediction in clinical settings where explanations must withstand clinical scrutiny.

This study has several limitations. First, the cohort comprises at-risk children from a single tertiary center (Helsinki University Hospital), which limits generalizability; both prevalence and predictor distributions may differ in other settings. Second, the observation window is restricted to the first 90 days of life, excluding later environmental factors, clinical events, and developmental assessments. Genetic data were largely unavailable, precluding assessment of heritable contributions. Third, maternal medication data could not be fully extracted from one of the source systems, meaning that maternal medication exposure may be incompletely represented in the model input. Fourth, the interpretability framework is predominantly univariate; although source-level and grouped phenobarbital ablation partially capture joint effects, the framework does not systematically evaluate variable interactions or higher-order dependencies. Fifth, the low outcome prevalence (8%) and etiological heterogeneity place inherent limits on attainable classification performance, and performance variability is summarized as mean *±* standard deviation across five cross-validation folds, which may not fully capture the uncertainty in model performance.

## Conclusion

Sequence-based modeling of longitudinal neonatal EHR data from the first 90 days of life can identify clinically meaningful signals for long-term neuropsychiatric risk, even when the outcomes are multifactorial and etiologically heterogeneous. The principal contribution of this work is methodological: a multi-method interpretability framework that provides internal consistency checks, distinguishing robust predictive signals from method-specific artifacts. By enabling clinicians to verify which model-derived insights are robust, this approach can strengthen the translation of predictive models into practice and provides a foundation for future condition-specific prediction, external validation, and risk stratification in neonatal follow-up care.

## Data availability statement

Due to Finnish data protection legislation and the EU General Data Protection Regulation, individual-level patient data cannot be made publicly available. Qualified researchers may apply for access to equivalent data through the HUS Data Service (https://www.hus.fi/en/research-and-education/tutkijan-palvelut/data-services). Studies combining HUS data with data from other controllers require a permit from Findata, the Social and Health Data Permit Authority (https://findata.fi/en/).

## Code availability

The STraTS model implementation is publicly available from the original authors [15] at https://github.com/sindhura97/STraTS. Logistic Regression and Random Forest were implemented using scikit-learn [45] (https://scikit-learn.org), and XGBoost using the XGBoost Python package (https://github.com/dmlc/xgboost). Analysis code is available from the corresponding author upon request.

## Acknowledgments

We gratefully recognize the contributions of Dr. Samuli Rautava, Dr. Marjo Metsäranta, and Dr. Matti Hero to this work. Moreover, we wish to acknowledge the support of Mona Halme, Mira Kilpi, and Mira Kuusinen from the Children and Adolescents’ Data Services Team at Helsinki University Hospital (HUS).

## Notes

### Competing Interest Statement

The authors have declared no competing interest.

### Funding Statement

This study was funded by The Foundation for Pediatric Research (Finland), the Association of Friends of the University Children's Hospitals (Lastenklinikoiden Kummit ry), and the Research Council of Finland (decision #359135).

### Author Declarations

Ethical approval and research permit were granted by Helsinki University Hospital, New Children's Hospital by Jari Petäjä, Director, Children and Adolescents (HUS/617/2025). According to Finnish legislation, Helsinki University Ethics Board only assesses clinical studies. Retrospective register studies, such as the current study, are approved by registry authorities, in this case Helsinki University Hospital, which assessed ethical concerns and data privacy issues and gave permission.

## References

1. Walter-Nicolet E, Marchand-Martin L, Morgan AS, Kaminski M, Benhammou V, Ancel PY, et al. Neurodevelopmental outcomes at five years in children born very preterm (24–31 weeks) exposed to opioids with or without midazolam: results from the French nationwide EPIPAGE-2 cohort study. The Lancet Regional Health–Europe. 2025;52.

2. Johnson S, Marlow N. Preterm birth and childhood psychiatric disorders. Pediatric research. 2011;69(8):11–8.

3. Yin W, Döring N, Persson MS, Persson M, Tedroff K, Ådén U, et al. Gestational age and risk of intellectual disability: a population-based cohort study. Archives of Disease in Childhood. 2022;107(9):826–32.

4. Crump C, Sundquist J, Sundquist K. Preterm or early term birth and risk of autism. Pediatrics. 2021;148(3):e2020032300.

5. Nosarti C, Reichenberg A, Murray RM, Cnattingius S, Lambe MP, Yin L, et al. Preterm birth and psychiatric disorders in young adult life. Archives of general psychiatry. 2012;69(6):610–7.

6. Anderson PJ, de Miranda DM, Albuquerque MR, Indredavik MS, Evensen KAI, Van Lieshout R, et al. Psychiatric disorders in individuals born very preterm/very low-birth weight: an individual participant data (IPD) meta-analysis. EClinicalMedicine. 2021;42.

7. Vanes LD, Murray RM, Nosarti C. Adult outcome of preterm birth: Implications for neurodevelopmental theories of psychosis. Schizophrenia Research. 2022;247:41–54.

8. Glass HC, Glidden D, Jeremy RJ, Barkovich AJ, Ferriero DM, Miller SP. Clinical neonatal seizures are independently associated with outcome in infants at risk for hypoxic-ischemic brain injury. The Journal of pediatrics. 2009;155(3):318–23.

9. Pisani F, Spagnoli C. Neonatal seizures: a review of outcomes and outcome predictors. Neuropediatrics. 2016;47(01):012–9.

10. Agrawal S, Rao SC, Bulsara MK, Patole SK. Prevalence of autism spectrum disorder in preterm infants: a meta-analysis. Pediatrics. 2018;142(3):e20180134.

11. Gardener H, Spiegelman D, Buka SL. Perinatal and neonatal risk factors for autism: a comprehensive meta-analysis. Pediatrics. 2011;128(2):344–55.

12. Harutyunyan H, Khachatrian H, Kale DC, Ver Steeg G, Galstyan A. Multitask learning and benchmarking with clinical time series data. Scientific data. 2019;6(1):96.

13. Lipton ZC, Kale DC, Elkan C, Wetzel R. Learning to diagnose with LSTM recurrent neural networks. In: International Conference on Learning Representations (ICLR); 2016. .

14. Baytas IM, Xiao C, Zhang X, Wang F, Jain AK, Zhou J. Patient subtyping via time-aware LSTM networks. In: Proceedings of the 23rd ACM SIGKDD international conference on knowledge discovery and data mining; 2017. p. 65-74.

15. Tipirneni S, Reddy CK. Self-supervised transformer for sparse and irregularly sampled multivariate clinical time-series. ACM Transactions on Knowledge Discovery from Data (TKDD). 2022;16(6):1–17.

16. Xu Y, Xu S, Ramprassad M, Tumanov A, Zhang C. TransEHR: self-supervised transformer for clinical time series data. In: Machine Learning for Health (ML4H). PMLR; 2023. p. 623–35.

17. Yang Z, Mitra A, Liu W, Berlowitz D, Yu H. TransformEHR: transformer-based encoder-decoder generative model to enhance prediction of disease outcomes using electronic health records. Nature communications. 2023;14(1):7857.

18. De Francesco D, Reiss JD, Roger J, Tang AS, Chang AL, Becker M, et al. Data-driven longitudinal characterization of neonatal health and morbidity. Science translational medicine. 2023;15(683):eadc9854.

19. Dick K, Kaczmarek E, Ducharme R, Bowie AC, Dingwall-Harvey AL, Howley H, et al. Transformer-based deep learning ensemble framework predicts autism spectrum disorder using health administrative and birth registry data. Scientific Reports. 2025;15(1):11816.

20. Rajagopalan SS, Tammimies K. Predicting neurodevelopmental disorders using machine learning models and electronic health records–status of the field. Journal of Neurodevelopmental Disorders. 2024;16(1):63.

21. Doshi-Velez F, Kim B. Considerations for evaluation and generalization in interpretable machine learning. In: Explainable and Interpretable Models in Computer Vision and Machine Learning. The Springer Series on Challenges in Machine Learning. Springer; 2018. p. 3–17.

22. Arrieta AB, Díaz-Rodríguez N, Del Ser J, Bennetot A, Tabik S, Barbado A, et al. Explainable Artificial Intelligence (XAI): Concepts, taxonomies, opportunities and challenges toward responsible AI. Information fusion. 2020;58:82–115.

23. Breiman L. Random Forests. Machine Learning. 2001;45(1):5–32. doi:10.1023/A:1010933404324.

24. Friedman JH. Greedy function approximation: a gradient boosting machine. Annals of statistics. 2001:1189–232.

25. Hosmer DW, Lemeshow S, Sturdivant RX. Applied Logistic Regression. 3rd ed. Wiley; 2013.

26. Chen T, Guestrin C. XGBoost: A Scalable Tree Boosting System. In: Proceedings of the 22nd ACM SIGKDD International Conference on Knowledge Discovery and Data Mining; 2016. p. 785–94.

27. Fisher A, Rudin C, Dominici F. All models are wrong, but many are useful: Learning a variable’s importance by studying an entire class of prediction models simultaneously. Journal of Machine Learning Research. 2019;20(177):1–81.

28. Covert I, Lundberg S, Lee SI. Explaining by removing: A unified framework for model explanation. Journal of Machine Learning Research. 2021;22(209):1–90.

29. Zeiler MD, Fergus R. Visualizing and understanding convolutional networks. In: European conference on computer vision. Springer; 2014. p. 818–33.

30. Lundberg SM, Lee SI. A unified approach to interpreting model predictions. Advances in neural information processing systems. 2017;30.

31. Saito T, Rehmsmeier M. The precision-recall plot is more informative than the ROC plot when evaluating binary classifiers on imbalanced datasets. PloS one. 2015;10(3):e0118432.

32. Polderman TJ, Benyamin B, de Leeuw CA, Sullivan PF, van Bochoven A, Visscher PM, et al. Meta-analysis of the heritability of human traits based on fifty years of twin studies. Nature Genetics. 2015;47(7):702–9. doi:10.1038/ng.3285.

33. Tick B, Bolton P, Happé F, Rutter M, Rijsdijk F. Heritability of autism spectrum disorders: a meta-analysis of twin studies. Journal of Child Psychology and Psychiatry. 2016;57(5):585–95. doi:10.1111/jcpp.12499.

34. Faraone SV, Asherson P, Banaschewski T, Biederman J, Buitelaar JK, Ramos-Quiroga JA, et al. Attention-deficit/hyperactivity disorder. Nature Reviews Disease Primers. 2015;1:15020. doi:10.1038/nrdp.2015.20.

35. Grieco J, Pulsifer M, Seligsohn K, Skotko B, Schwartz A. Down syndrome: cognitive and behavioral functioning across the lifespan. American Journal of Medical Genetics Part C: Seminars in Medical Genetics. 2015;169(2):135–49. doi:10.1002/ajmg.c.31439.

36. Linsell L, Malouf R, Morris J, Kurinczuk JJ, Marlow N. Prognostic factors for poor cognitive development in children born very preterm or with very low birth weight: a systematic review. JAMA Pediatrics. 2015;169(12):1162–72. doi:10.1001/jamapediatrics.2015.2175.

37. Loomes R, Hull L, Mandy WPL. What is the male-to-female ratio in autism spectrum disorder? A systematic review and meta-analysis. Journal of the American Academy of Child & Adolescent Psychiatry. 2017;56(6):466–74. doi:10.1016/j.jaac.2017.03.013.

38. Subedi D, DeBoer MD, Scharf RJ. Developmental trajectories in children with prolonged NICU stays. Archives of Disease in Childhood. 2017;102(1):29–34. doi:10.1136/archdischild-2016-310777.

39. Kumar IE, Venkatasubramanian S, Scheidegger C, Friedler S. Problems with Shapley-value-based explanations as feature importance measures. In: International conference on machine learning. PMLR; 2020. p. 5491–500.

40. Huang X, Marques-Silva J. On the failings of Shapley values for explainability. International Journal of Approximate Reasoning. 2024;171:109112.

41. Ghassemi M, Oakden-Rayner L, Beam AL. The false hope of current approaches to explainable artificial intelligence in health care. The Lancet Digital Health. 2021;3(11):e745–50. doi:10.1016/S2589-7500(21)00208-9.

42. Bowe AK, Lightbody G, Staines A, Murray DM, Norman M. Prediction of 2-year cognitive outcomes in very preterm infants using machine learning methods. JAMA Network Open. 2023;6(12):e2349111–1.

43. Bowe AK, Lightbody G, O’Boyle DS, Staines A, Murray DM. Predicting low cognitive ability at age 5 years using perinatal data and machine learning. Pediatric Research. 2024;95(6):1634–43.

44. Ortega-Leon A, Urda D, Turias IJ, Lubían-López SP, Benavente-Fernández I. Machine learning techniques for predicting neurodevelopmental impairments in premature infants: a systematic review. Frontiers in artificial intelligence. 2025;8:1481338.

45. Pedregosa F, Varoquaux G, Gramfort A, Michel V, Thirion B, Grisel O, et al. Scikit-learn: Machine Learning in Python. Journal of Machine Learning Research. 2011;12:2825–30.

