## Supplementary material for "Predicting long-term adverse outcomes after neonatal intensive care": S5 Table

**Table S5: Important variables identified by perturbation-based importance and LOO feature attribution analyses.**

| **Variable** | **N** | **Perturb.** | **LOO** |
|---|---|---|---|
| *Identified by both methods* | | | |
| Birth weight | 17,655 | ✓ | ✓ |
| Gender | 17,655 | ✓ | ✓ |
| Apgar (1 min) | 17,655 | ✓ | ✓ |
| uS-TSH | 17,469 | ✓ | ✓ |
| Treatment time in hospital (days) | 17,622 | ✓ | ✓ |
| *Identified by perturbation only* | | | |
| Chromosomal abn. (Q90) | 103 | ✓ | |
| Other disturbances of cerebral status of newborn (P91) | 217 | ✓ | |
| Encounters (exams/invest.) (Z01) | 1,569 | ✓ | |
| Congenital Malformations of Cardiac Septa (Q21) | 718 | ✓ | |
| Phenobarbital | 202 | ✓ | |
| Phenobarbital (serum) | 193 | ✓ | |
| Lupus antibody | 74 | ✓ | |
| Creatinine | 2,637 | ✓ | |
| Cong. malf. (Q87) | 78 | ✓ | |
| RDW | 15,456 | ✓ | |
| uA-BE | 16,459 | ✓ | |
| Levetiracetam | 63 | ✓ | |
| *Identified by LOO only* | | | |
| Gestational age (days) | 17,655 | | ✓ |
| Delivery type | 17,655 | | ✓ |
| Birth length | 16,768 | | ✓ |
| Head circumference | 15,120 | | ✓ |
| No. of fetuses | 17,655 | | ✓ |
| Apgar (5 min) | 12,479 | | ✓ |
| Apgar (10 min) | 8,920 | | ✓ |
| Ua-pO2 | 16,457 | | ✓ |
| cB-pO2 | 13,201 | | ✓ |
| E-MCHC | 15,792 | | ✓ |
| E-MCV | 15,792 | | ✓ |
| B-HKR | 15,875 | | ✓ |
| B-Trom | 15,787 | | ✓ |
| P-Ca-IonA | 14,434 | | ✓ |
| BMI (growth) | 16,841 | | ✓ |

*N* denotes the number of children with the variable observed (non-missing or present).
