## Supplementary material for "Predicting long-term adverse outcomes after neonatal intensive care": S4 Table

**Table S4: Hyperparameter optimization summary and search spaces (all models).**

| Model | Hyperparameter | Values / range | Notes |
|---|---|---|---|
| **Logistic Regression** | Solver | `lbfgs`, `liblinear`, `saga` | Solver–penalty compatibility enforced |
| | Penalty | `l1`, `l2`, `elasticnet` | As supported by solver |
| | Regularization strength ($C$) | $10^{-4}$ to $10^{2}$ (log-uniform) | |
| | Elastic Net mixing (`l1_ratio`) | {0.1, 0.5, 0.9} | Elastic Net only (typically `saga`) |
| | Max iterations (`max_iter`) | {300, 500, 800} | |
| | Class weighting (`class_weight`) | `balanced` | Address class imbalance |
| | **# configurations evaluated** | **20** | Random search |
| **XGBoost** | `n_estimators` | 50–1000 | Boosting rounds |
| | `max_depth` | 3–15 | |
| | `learning_rate` | 0.01–0.3 | |
| | `subsample` | 0.6–1.0 | Row subsampling |
| | `colsample_bytree` | 0.6–1.0 | Column subsampling (tree) |
| | `colsample_bylevel` | 0.6–1.0 | Column subsampling (level) |
| | `reg_alpha` | 0–10 | L1 regularization |
| | `reg_lambda` | 0–10 | L2 regularization |
| | `min_child_weight` | 1–10 | |
| | `gamma` | 0–5 | Minimum loss reduction |
| | **# configurations evaluated** | **20** | Random search |
| **Random Forest** | `n_estimators` | 50–500 | |
| | `max_depth` | `None` or 3–20 | |
| | `min_samples_split` | 2–20 | |
| | `min_samples_leaf` | 1–10 | |
| | `max_features` | `sqrt`, `log2`, 0.5, 0.7, 0.8, `all` | Feature subsampling |
| | `bootstrap` | `True` / `False` | |
| | `class_weight` | `balanced`, `balanced_subsample`, `none` | |
| | **# configurations evaluated** | **20** | Random search |
| **STraTS** | Embedding dimension | {8, 16, 32} | Latent embedding size |
| | # Transformer layers | {2, 3, 4} | |
| | # Attention heads | {8, 12} | |
| | Feed-forward dropout | {0.0, 0.1} | |
| | Attention dropout | {0.0, 0.1} | |
| | Learning rate | $10^{-4}$–$5 \times 10^{-4}$ | |
| | Batch size | {8, 16, 32} | Evaluation batch size matched to training |
| | **# configurations evaluated** | **20** | Random search |

Hyperparameters were tuned via random search. For each model, 20 hyperparameter configurations were evaluated.
