## Supplementary material for "Predicting long-term adverse outcomes after neonatal intensive care": S3 Table

**Table S3: ICD-10 code blocks treated as repeatable diagnoses and retained at monthly resolution.**

| ICD-10 block (3-char) | Included categories |
|---|---|
| A00–A09 | A00, A01, . . . , A09 |
| B00–B99 | B00, B01, . . . , B99 |
| G00–G09 | G00, G01, . . . , G09 |
| H10 | H10 (incl. H10.x) |
| J00–J22 | J00, J01, . . . , J22 |
| K35–K38 | K35, K36, K37, K38 |
| K65 | K65 (incl. K65.x) |
| L00–L08 | L00, L01, . . . , L08 |
| N10 | N10 |
| N30 | N30 |
| N39 | N39 |
| S00–S99 | S00, S01, . . . , S99 |
| T00–T99 | T00, T01, . . . , T99 |

ICD-10 codes were truncated to the 3-character category (decimal part removed). Diagnoses falling within the listed blocks were modeled as potentially recurring events and therefore kept as month-specific occurrences rather than being collapsed into a single ever/never indicator.
