## Supplementary material for "Predicting long-term adverse outcomes after neonatal intensive care": S2 Table

**Table S2: List of medications for mothers and children.**

| Neonatal ICU (child) | | | Maternal (pregnancy) | |
|---|---|---|---|---|
| Drug | ATC code | Dose unit | Drug | ATC code |
| Glucocorticoids (inhaled antiinfectives) | R03BA | mg | Dexamethasone | H02AB02 |
| Dexamethasone | H02AB02 | mg | Betamethasone | H02AB01 |
| Hydrocortisone | H02AB09 | mg | Antiepileptics | N03A |
| Insulin (human) | A10AB01 | IU | Antipsychotics | N05A |
| Insulin aspart | A10AB05 | IU | Anxiolytics | N05B |
| Natural phospholipids | R07AA02 | mg | Hypnotics and sedatives | N05C |
| Caffeine | N06BC01 | mg | Antidepressants | N06A |
| Budesonide | R03BA02 | mg | Psychostimulants | N06B |
| Ibuprofen | C01EB16 | mg | Drugs used in addictive disorders | N07B |
| Indometacin | C01EB03 | mg | Psycholeptics and psychoanaleptics (comb.) | N06C |
| Morphine | N02AA01 | mg | | |
| Fentanyl | N02AB03 | mg | | |
| Ketamine | N01AX03 | mg | | |
| Dexmedetomidine | N05CM18 | mg | | |
| Midazolam | N05CD08 | mg | | |
| Lorazepam | N05BA06 | mg | | |
| Phenobarbital | N03AA02 | mg | | |
| Fosphenytoin | N03AB05 | mg FE | | |
| Levetiracetam | N03AX14 | mg | | |

Medications included in the study, their ATC codes, and dose units. Neonatal medication doses are recorded as daily cumulative dose per kg body weight. Maternal medications are encoded as binary presence during pregnancy.
