## Supplementary material for "Predicting long-term adverse outcomes after neonatal intensive care": S1 Table

**Table S1: Extended dataset variable summary.**

| Variable | Type | Children No MND (occ.) | Children MND (occ.) | No MND Mean/Median | MND Mean/Median | Adj. $p_{FDR}$ |
|---|---|---|---|---|---|---|
| **Demographics** | | | | | | |
| Birth weight (gr) | Continuous | 16235 (16235) | 1420 (1420) | 3131.53 (878.15) | 2898.84 (1014.14) | 7.01e-20* |
| Gender | Categorical (1-2) | 16235 (16235) | 1420 (1420) | 2.0 | 2.0 | |
| Apgar 1min | Ordinal (1-10) | 16032 (16032) | 1400 (1400) | 9.0 | 9.0 | |
| Apgar 5min | Ordinal (1-10) | 11441 (11441) | 1038 (1038) | 9.0 | 9.0 | |
| Apgar 10min | Ordinal (1-10) | 8175 (8175) | 745 (745) | 10.0 | 10.0 | |
| Number Of Fetuses | Ordinal (1-4) | 16235 (16235) | 1420 (1420) | 1.0 | 1.0 | |
| Delivery Type | Categorical (1-4) | 16235 (16235) | 1420 (1420) | 1.0 | 1.0 | |
| Birth length (cm) | Continuous | 15442 (15442) | 1326 (1326) | 48.51 (4.19) | 47.26 (5.16) | 3.36e-23* |
| Head circumference at birth (cm) | Continuous | 13995 (13995) | 1125 (1125) | 34.20 (2.53) | 33.83 (2.81) | 1.59e-05* |
| Gestational age (days) | Continuous | 16235 (16235) | 1420 (1420) | 266.20 (23.37) | 260.39 (28.92) | 2.54e-17* |
| **Growth** | | | | | | |
| BMI | Continuous | 15505 (31069) | 1336 (3875) | 12.78 (2.29) | 12.66 (2.27) | 0.007* |
| Height (cm) | Continuous | 15505 (30881) | 1336 (3853) | 48.12 (5.21) | 47.91 (5.89) | 0.055 |
| Weight (kg) | Continuous | 15830 (68302) | 1376 (9281) | 2.86 (1.11) | 2.89 (1.14) | 0.063 |
| **Hospital** | | | | | | |
| Treatment time in hospital (days) | Presence | 16220 (158542) | 1420 (24550) | 99.91% | 100.00% | 0.748 |
| **Lab** | | | | | | |
| S-Fenob (umol/l) | Continuous | 121 (348) | 72 (322) | 103.02 (28.36) | 102.43 (29.20) | 0.923 |
| uS-TSH (mU/l) | Continuous | 16092 (16095) | 1396 (1396) | 8.39 (10.01) | 9.41 (29.62) | 0.017* |
| P-Krea (umol/l) | Continuous | 2234 (6957) | 421 (1687) | 56.44 (44.73) | 57.07 (43.27) | 0.788 |
| P-LupusAK (-) | Binary | 43 (44) | 32 (32) | 0.00 | 0.00 | 0.977 |
| uA-Be (mmol/l) | Continuous | 15163 (15163) | 1296 (1296) | -4.65 (3.85) | -4.58 (4.06) | 0.748 |
| uA-pO2 (kPa) | Continuous | 15162 (15162) | 1295 (1295) | 2.74 (1.17) | 2.74 (1.20) | 1.000 |
| E-RDW (%) | Continuous | 14171 (84337) | 1285 (13350) | 17.13 (2.66) | 17.64 (2.51) | 4.89e-97* |
| uA-pH (-) | Continuous | 15210 (15210) | 1299 (1299) | 7.23 (0.10) | 7.24 (0.11) | 0.479 |
| uA-HCO3-St (mmol/l) | Continuous | 15132 (15132) | 1289 (1289) | 18.86 (2.97) | 18.93 (2.99) | 0.631 |
| cB-pH (-) | Continuous | 13230 (84108) | 1223 (13468) | 7.36 (0.05) | 7.36 (0.05) | 1.22e-12* |

Continued on next page

**Table S1:** Dataset variable summary (continued).

| Variable | Type | Children No MND (occ.) | Children MND (occ.) | No MND Mean/Median | MND Mean/Median | Adj. $p_{FDR}$ |
|---|---|---|---|---|---|---|
| fP-NH4-ion (umol/l) | Continuous | 278 (401) | 84 (166) | 55.22 (23.33) | 60.80 (23.17) | 0.034* |
| cB-Bil (umol/l) | Continuous | 2640 (10399) | 240 (981) | 148.86 (62.93) | 149.60 (66.10) | 0.881 |
| P -AFOS (IU/l) | Continuous | 2348 (6377) | 347 (1125) | 324.01 (114.60) | 336.66 (119.16) | 0.003* |
| cB-HCO3-St (mmol/l) | Continuous | 13224 (83919) | 1222 (13440) | 24.54 (2.53) | 25.07 (2.89) | 1.16e-105* |
| uA-pCO2 (kPa) | Continuous | 15179 (15179) | 1297 (1297) | 7.59 (1.75) | 7.54 (1.84) | 0.527 |
| **Child medications** | | | | | | |
| Phenobarbital (mg/kg/day) | Continuous | 126 (430) | 76 (401) | 9.08 (8.04) | 7.79 (7.23) | 0.052 |
| Levetiracetam (mg/kg/day) | Continuous | 33 (132) | 30 (274) | 21.44 (10.28) | 28.55 (15.08) | 9.27e-06* |
| Midazolam (mg/kg/day) | Continuous | 175 (852) | 43 (204) | 1.23 (1.13) | 1.77 (1.40) | 3.68e-08* |
| Insulin Human (IU/kg/day) | Continuous | 159 (911) | 62 (389) | 0.79 (0.95) | 1.08 (1.70) | 3.68e-04* |
| Ketamine (mg/kg/day) | Continuous | 849 (1782) | 181 (437) | 4.18 (7.49) | 6.09 (13.00) | 2.94e-04* |
| Dexamethasone (mg/kg/day) | Continuous | 119 (777) | 30 (172) | 0.27 (0.40) | 0.32 (0.46) | 0.328 |
| Fentanyl (mg/kg/day) | Continuous | 741 (3167) | 159 (1035) | 0.02 (0.01) | 0.02 (0.02) | 0.436 |
| Ibuprofen (mg/kg/day) | Continuous | 40 (89) | 7 (20) | 8.50 (4.84) | 6.94 (2.43) | 0.358 |
| Dexmedetomidine (mg/kg/day) | Continuous | 186 (1218) | 36 (266) | 0.01 (0.01) | 0.01 (0.01) | 0.075 |
| Lorazepam (mg/kg/day) | Continuous | 31 (147) | 5 (22) | 0.27 (0.25) | 0.38 (0.27) | 0.120 |
| Glucocorticoids Inhaled Antiinfectives (mg/kg/day) | Continuous | 43 (594) | 12 (150) | 0.25 (0.16) | 0.29 (0.16) | 0.030* |
| Morphine (mg/kg/day) | Continuous | 523 (4151) | 86 (618) | 0.41 (0.39) | 0.40 (0.35) | 0.843 |
| Insulin Aspart (IU/kg/day) | Continuous | 84 (169) | 12 (21) | 0.35 (0.32) | 0.34 (0.31) | 0.953 |
| Natural Phospholipids (mg/kg/day) | Continuous | 577 (721) | 103 (127) | 146.08 (52.93) | 161.42 (68.46) | 0.017* |
| Indometacin (mg/kg/day) | Continuous | 116 (245) | 28 (57) | 0.18 (0.07) | 0.18 (0.06) | 1.000 |
| Hydrocortisone (mg/kg/day) | Continuous | 252 (3884) | 71 (995) | 2.16 (1.52) | 2.36 (1.46) | 6.73e-04* |
| Caffeine (mg/kg/day) | Continuous | 1353 (29892) | 196 (5910) | 7.25 (3.59) | 7.20 (3.18) | 0.585 |
| **Diagnoses** | | | | | | |
| Chromosomal Abnormalities Not Elsewhere Classified (Q90) | Presence | 47 (70) | 58 (82) | 0.29% | 4.08% | 3.12e-68* |
| Other disturbances of cerebral status of newborn (P91) | Presence | 147 (168) | 75 (88) | 0.91% | 5.28% | 1.75e-43* |

Continued on next page

**Table S1:** Dataset variable summary (continued).

| Variable | Type | Children No MND (occ.) | Children MND (occ.) | No MND Mean/Median | MND Mean/Median | Adj. $p_{FDR}$ |
|---|---|---|---|---|---|---|
| Encounters For Examinations Investigations (Z01) | Presence | 1608 (1753) | 313 (358) | 9.90% | 22.04% | 2.41e-43* |
| Congenital malformation syndromes (Q87) | Presence | 48 (51) | 30 (35) | 0.30% | 2.11% | 6.25e-21* |
| Congenital Malformations of Cardiac Septa (Q21) | Presence | 641 (871) | 128 (180) | 3.95% | 9.01% | 8.69e-18* |
| Specific Health Care Encounters (Z50) | Presence | 534 (535) | 120 (120) | 3.29% | 8.45% | 2.20e-21* |
| Transitory Endocrine Metabolic Disorders Newborn (P70) | Presence | 4104 (4305) | 449 (469) | 25.28% | 31.62% | 1.39e-06* |
| Behavioral Syndromes Associated With Physiological Disturbances (F59) | Presence | 20 (20) | 13 (13) | 0.12% | 0.92% | 2.58e-09* |
| Congenital Malformations Eye Ear Face Neck (Q17) | Presence | 55 (58) | 19 (22) | 0.34% | 1.34% | 5.70e-07* |
| Perinatal Respiratory Cardiovascular Disorders (P21) | Presence | 1350 (1420) | 128 (140) | 8.32% | 9.01% | 0.630 |
| **Maternal medication** | | | | | | |
| Anxiolytics | Presence | 1472 (5235) | 86 (334) | 9.07% | 6.06% | 7.13e-04* |
| Antiepileptics | Presence | 369 (2470) | 48 (327) | 2.27% | 3.38% | 0.038* |
| Psychostimulants | Presence | 16 (125) | <3 (11) | 0.10% | - | - |
| Used Addictive Disorders | Presence | 97 (567) | 7 (35) | 0.60% | 0.49% | 0.902 |
| Hypnotics Sedatives | Presence | 217 (1340) | 16 (90) | 1.34% | 1.13% | 0.783 |
| Psycholeptics Psychoanaleptics Combination | Presence | 8 (60) | <3 (20) | 0.05% | - | - |
| Dexamethasone | Presence | 18 (67) | <3 (4) | 0.11% | - | - |
| Antipsychotics | Presence | 521 (3258) | 39 (194) | 3.21% | 2.75% | 0.627 |
| Betamethasone | Presence | 34 (71) | 5 (11) | 0.21% | 0.35% | 0.663 |
| Antidepressants | Presence | 836 (5100) | 72 (387) | 5.15% | 5.07% | 1.000 |

* indicates FDR-adjusted $p < 0.05$ (Benjamini-Hochberg).
