## Supplementary material for "Predicting long-term adverse outcomes after neonatal intensive care": S1 Fig

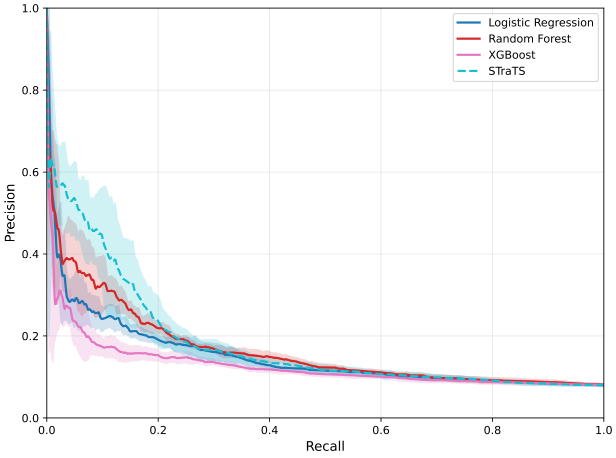


**(A)**

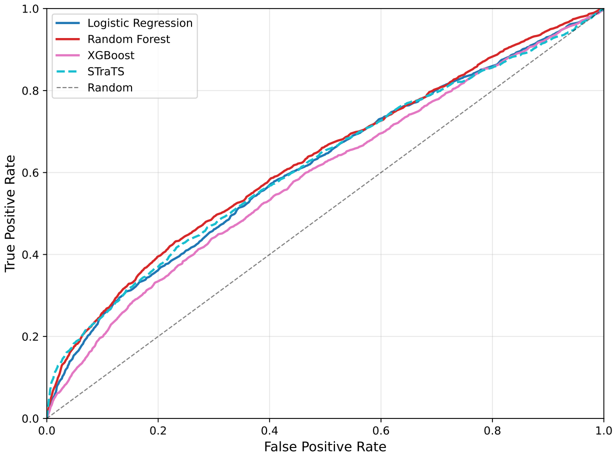


**(B)**

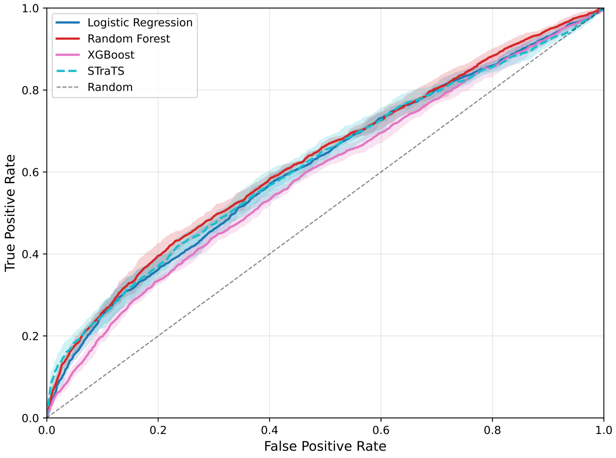


**(C)**

**Figure S1: Precision–recall and ROC curves with cross-fold variability.** (A) Precision–recall curves across folds. (B) Mean ROC curves. (C) ROC curves across folds.
